# Physical activity and cardiometabolic health across an extreme lifestyle gradient

**DOI:** 10.1101/2025.07.11.25331394

**Authors:** Thomas S. Kraft, Ian J. Wallace, Yvonne A L Lim, Kar Lye Tam, Tan Bee Ting A/P Tan Boon Huat, Rob Tennyson, Steven K. W. Chow, Kamal Solhaimi bin Fadzil, Colin Nicholas, Izandis bin Mohd Sayed, Vivek V. Venkataraman, Amanda J. Lea

## Abstract

Cardiometabolic diseases commonly rise with market-integration, industrialization, and urbanization, yet the direct role of physical activity remains debated. We collected accelerometry, health, and interview data from 1075 Orang Asli adults in Peninsular Malaysia, who currently span a wide range from rural, non-industrial to urban, industrialized lifestyles. More urban lifestyles were associated with reduced physical activity and worse cardiometabolic health. However, physical activity mediated only a small portion of urbanization’s effects, suggesting independent influences. Age-related declines in activity were steeper in urban settings, indicating that non-industrial lifestyles promote activity in old age and may buffer older adults from functional decline. These results support the view that declining physical activity is an evolutionarily novel challenge contributing to poor health in modern environments.

**Teaser:** Shifts toward more urban, market-integrated lifestyles are associated with lower levels of physical activity and worse cardiometabolic health, but physical activity explains only a small part of the lifestyle–health relationship.

## Introduction

The generally robust cardiometabolic health observed among non-industrial human societies stands in stark contrast to the escalating prevalence of obesity, type 2 diabetes, and cardiovascular disease among industrialized populations (*1*). These health disorders are notably rare or absent among many hunter-gatherer, pastoralist, and subsistence farming groups; further, cardiometabolic health frequently declines following transitions from non-industrial to more urban, market-integrated, and industrialized lifestyles (*2–5*). Together, these observations imply that elements of non-industrial lifestyles may offer protective benefits (*6–10*). In particular, it is commonly proposed that the characteristically high physical activity levels of non-industrial societies play a major role in preserving cardiometabolic health (*7*, *9–11*).

Although it is reasonable to expect that high levels of physical activity promote favorable cardiometabolic health profiles among non-industrial societies, multiple lines of evidence suggest that the benefits could be less than commonly assumed. First, studies using doubly labeled water have revealed similar levels of total daily energy expenditure between non-industrial and industrialized populations, suggesting that differences in dietary energy intake—rather than physical activity—primarily account for the variation in obesity risk between such populations (*12*, *13*). Second, studies of certain non-industrial societies have observed deterioration in cardiometabolic health among traditional-living people who have recently transitioned to more urban, industrialized environments or are undergoing some degree of modernization, despite them not exhibiting concomitant major declines in physical activity levels (*14–16*). Third, studies show that while non-industrial societies are highly active, their movement patterns consist primarily of light-to-moderate intensity activities such as walking and food processing (*17*, *18*). Though such activity has been shown to provide some cardiometabolic benefits in industrialized contexts, its effects are inconsistent and evidently smaller than the benefits of vigorous exercise (*19*, *20*). Finally, a previous study of physical activity (using primarily behavioral observations) and BMI, fat free mass, and percent body fat in a non-industrial population undergoing a relatively mild lifestyle transition failed to identify substantial effects (*14*).

The degree to which high physical activity levels in non-industrial societies explain their favorable cardiometabolic health remains debated, largely because there have been few studies of sufficient sample size directly linking objective measures of physical activity with cardiometabolic health across non-industrial and industrialized contexts (*9*, *14*). To address this gap, we investigate relationships between physical activity, cardiometabolic health biomarkers, and lifestyle variation among the Indigenous peoples of Peninsular Malaysia known as the Orang Asli. The Orang Asli are traditionally rainforest-dwelling hunter-gatherers, fishers, and subsistence farmers. However, in recent years, many individuals have experienced varying degrees of lifestyle change due to factors such as acculturation, national economic development, the expansion of built infrastructure, and integration into the market economy, with the result that today Orang Asli lifestyles span a wide spectrum from traditional, non-industrial to urban, industrial. Previous work by our group and others has shown that cardiometabolic health varies in a predictable way across this lifestyle spectrum, with the best health observed among the most traditional and least urbanized Orang Asli (*2*, *21*, *22*). Yet, the role of physical activity in shaping this health variation has not been established.

In this study, we collected data with 1075 Orang Asli individuals spanning an extreme lifestyle gradient to test three general hypotheses regarding physical activity. First, we used detailed ethnographic information on lifestyle variation and gold-standard accelerometry methods to provide detailed characterizations of physical activity patterns among a traditionally non-industrial population undergoing lifestyle change. We test the hypothesis that greater urbanization is associated with decreased levels of physical activity, examining the effects of aging on physical activity and how different types of activity (i.e., inactivity versus light, moderate, or vigorous activity) change with urbanization. Second, we collected high-dimensional cardiometabolic health biomarker data to test the hypothesis that physical activity levels are directly associated with better health outcomes at the individual level. Third, we used mediation analysis to assess the degree to which physical activity mediates the relationship between urbanization and cardiometabolic health.

## Results

As part of a long-term study of Orang Asli lifestyle, culture, and health—the Orang Asli Health and Lifeways project (*23*)—we collected wrist-worn accelerometry data using Axivity AX3 devices. Following quality control checks, physical activity information was available from 1075 participants (age range: 18 to 91; 63.6% female) for a total of 5892 valid days of activity (median = 6 days per person). We extracted eight standard physical activity metrics using open-source GGIR software (*24*)(Table 1), and observed generally high correlations among them (absolute average Pearson’s *r* = 0.42, range = 0.03 - 0.96), with some variables (e.g., time spent in moderate activity) showing high redundancy and others (e.g., intensity gradient) evincing only modest correlation to other metrics (Fig. 1A). As expected, time spent inactive was negatively correlated with other metrics, for which higher values represent greater amounts of activity. Based on these correlations, we identified three relatively independent metrics, representing theoretically different aspects of physical activity, as the focus of our main analysis: daily step counts (steps/day), time spent inactive (minutes/day), and intensity gradient (slope) (Fig. 1; analyses of the full suite of metrics included in Supplementary Materials).

**Figure 1:**
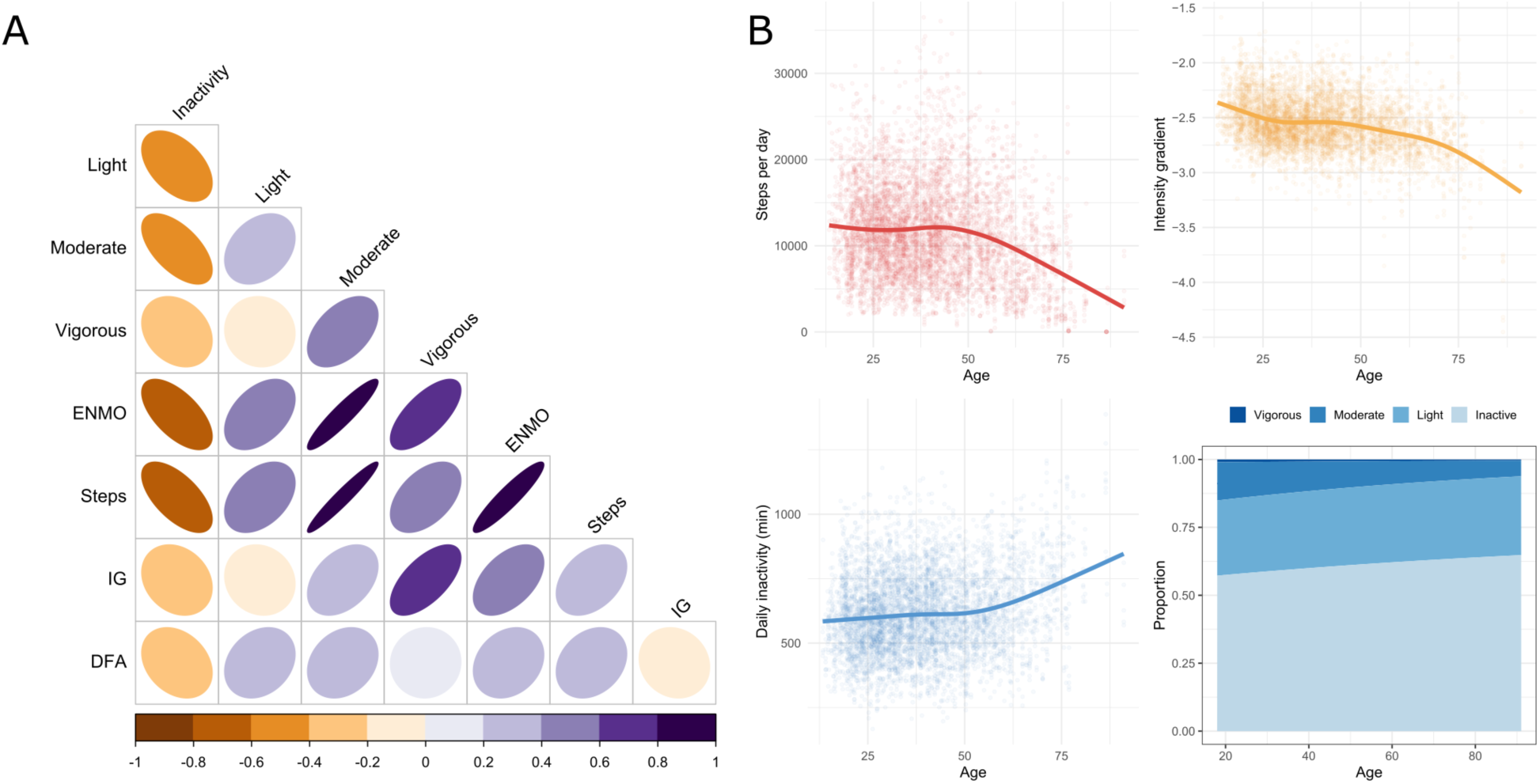
Physical activity and health outcomes in this study. (A) Correlations between different physical activity metrics extracted from accelerometry data (all correlations at the daily level except for DFA, which is a person-level metric). (B) Relationships of selected physical activity metrics with age. Smoothing curves are from generalized additive mixed models of physical activity outcomes as a function of a thin-plate spline for age and a random intercept for repeat observations of individuals (each observation is a single day of physical activity). Proportions of time spent in inactivity or vigorous/moderate/light activity as a function of age are derived from a multilevel dirichlet regression with a random intercept for repeat observations of individuals.

**Table 1:**
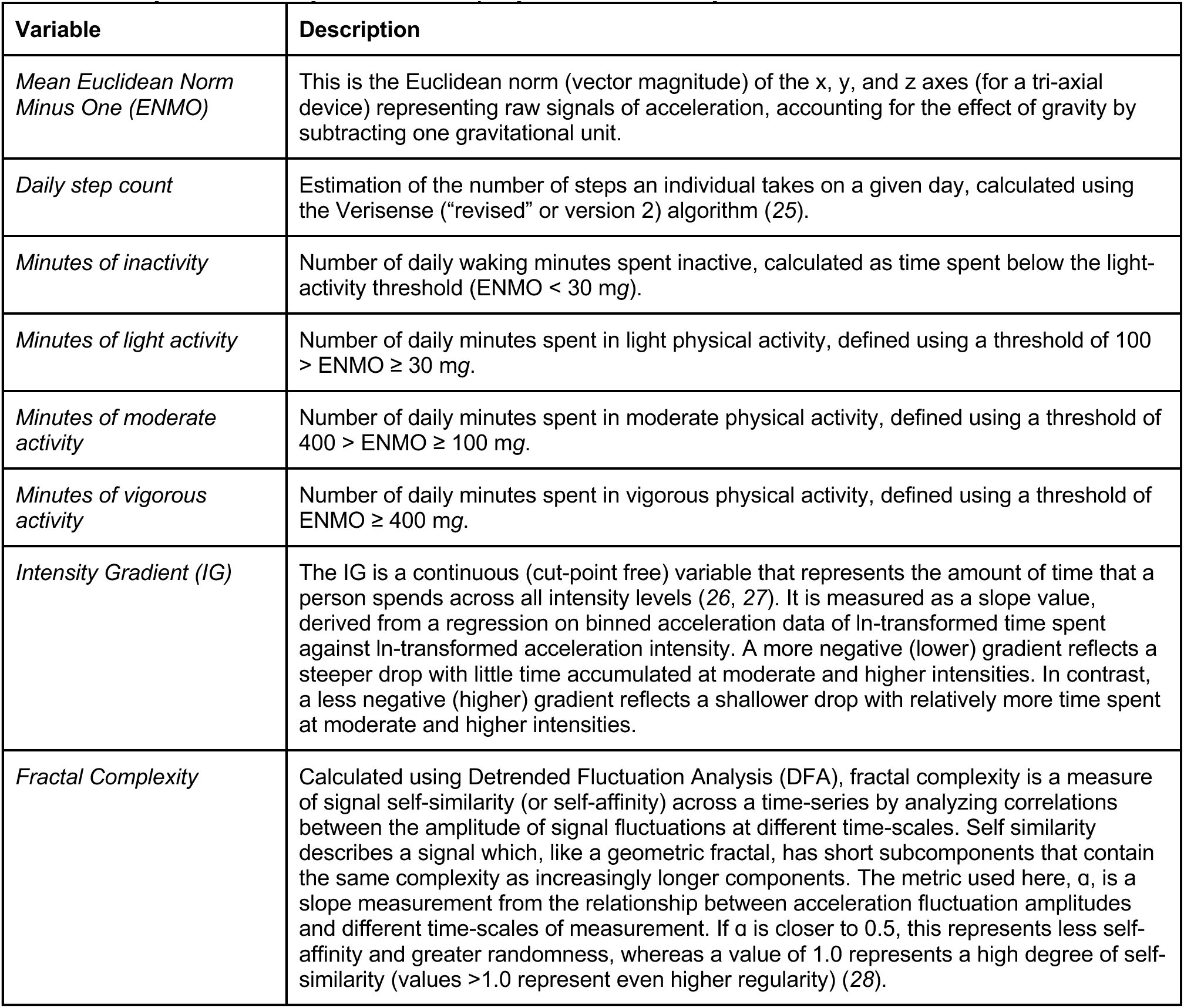
Physical activity metrics employed in this study.

| **Variable** | **Description** |
| --- | --- |
| *Mean Euclidean Norm Minus One (ENMO)* | This is the Euclidean norm (vector magnitude) of the x, y, and z axes (for a tri-axial device) representing raw signals of acceleration, accounting for the effect of gravity by subtracting one gravitational unit. |
| *Daily step count* | Estimation of the number of steps an individual takes on a given day, calculated using the Verisense ("revised" or version 2) algorithm (25). |
| *Minutes of inactivity* | Number of daily waking minutes spent inactive, calculated as time spent below the light-activity threshold ($ENMO < 30$ mg). |
| *Minutes of light activity* | Number of daily minutes spent in light physical activity, defined using a threshold of $100 > ENMO \geq 30$ mg. |
| *Minutes of moderate activity* | Number of daily minutes spent in moderate physical activity, defined using a threshold of $400 > ENMO \geq 100$ mg. |
| *Minutes of vigorous activity* | Number of daily minutes spent in vigorous physical activity, defined using a threshold of $ENMO \geq 400$ mg. |
| *Intensity Gradient (IG)* | The IG is a continuous (cut-point free) variable that represents the amount of time that a person spends across all intensity levels (26, 27). It is measured as a slope value, derived from a regression on binned acceleration data of $\ln$-transformed time spent against $\ln$-transformed acceleration intensity. A more negative (lower) gradient reflects a steeper drop with little time accumulated at moderate and higher intensities. In contrast, a less negative (higher) gradient reflects a shallower drop with relatively more time spent at moderate and higher intensities. |
| *Fractal Complexity* | Calculated using Detrended Fluctuation Analysis (DFA), fractal complexity is a measure of signal self-similarity (or self-affinity) across a time-series by analyzing correlations between the amplitude of signal fluctuations at different time-scales. Self similarity describes a signal which, like a geometric fractal, has short subcomponents that contain the same complexity as increasingly longer components. The metric used here, $\alpha$, is a slope measurement from the relationship between acceleration fluctuation amplitudes and different time-scales of measurement. If $\alpha$ is closer to 0.5, this represents less self-affinity and greater randomness, whereas a value of 1.0 represents a high degree of self-similarity (values $>1.0$ represent even higher regularity) (28). |

### Physical activity declines with urbanization

We also conducted detailed questionnaires with participants to ascertain information about salient lifestyle factors related to urbanization, industrialization, and market integration. These measures, specifically those focused on community-level attributes of the built environment and modernization like electricity access, water source, population density, wage labor engagement, and average education levels, were then combined into a previously described “urbanicity score.” This score has been demonstrated to effectively capture lifestyle variation in multiple non-industrial populations undergoing lifestyle change (*22*), and to predict cardiometabolic health in the Orang Asli specifically Importantly, this metric therefore captures a continuous range of lifestyle variation that is distinct from dichotomous epidemiological studies comparing urban and rural environments (e.g., *24*), and has already been well-validated and described for the study group.

As expected based on the extreme lifestyle gradient across Orang Asli communities, physical activity metrics varied substantially by location, ranging from villages with median daily step counts of ∼15,000 to 5,000 (Fig. 2A-B). In support of our first hypothesis—that greater urbanization would be associated with decreased levels of physical activity—we found that physical activity decreased as a function of urbanicity, with more urbanized individuals exhibiting significantly lower average step counts and intensity gradients, and greater time spent inactive (Fig. 2C). Differences between the most and least urbanized individuals ranged from an ∼0.75 standard deviation change for intensity gradient and time spent inactive, to an ∼0.3 change in daily step counts. The effects of urbanicity on declines in intensity gradient and increases in time spent inactive were thus stronger than observed trends for daily step counts, suggesting that lifestyle change is especially discouraging of higher intensity activities and instead encourages more inactivity. Male participants exhibited greater daily step counts and intensity gradient despite spending more time inactive, reflecting higher levels of moderate and vigorous physical activity (Fig. 2D; ꞵ [95% CI] = 0.08 [-0.02, 0.17], 0.49 [0.40, 0.59], 0.23 [0.14, 0.33] in models of standardized step counts, intensity gradient, and time spent inactive, respectively).

**Figure 2:**
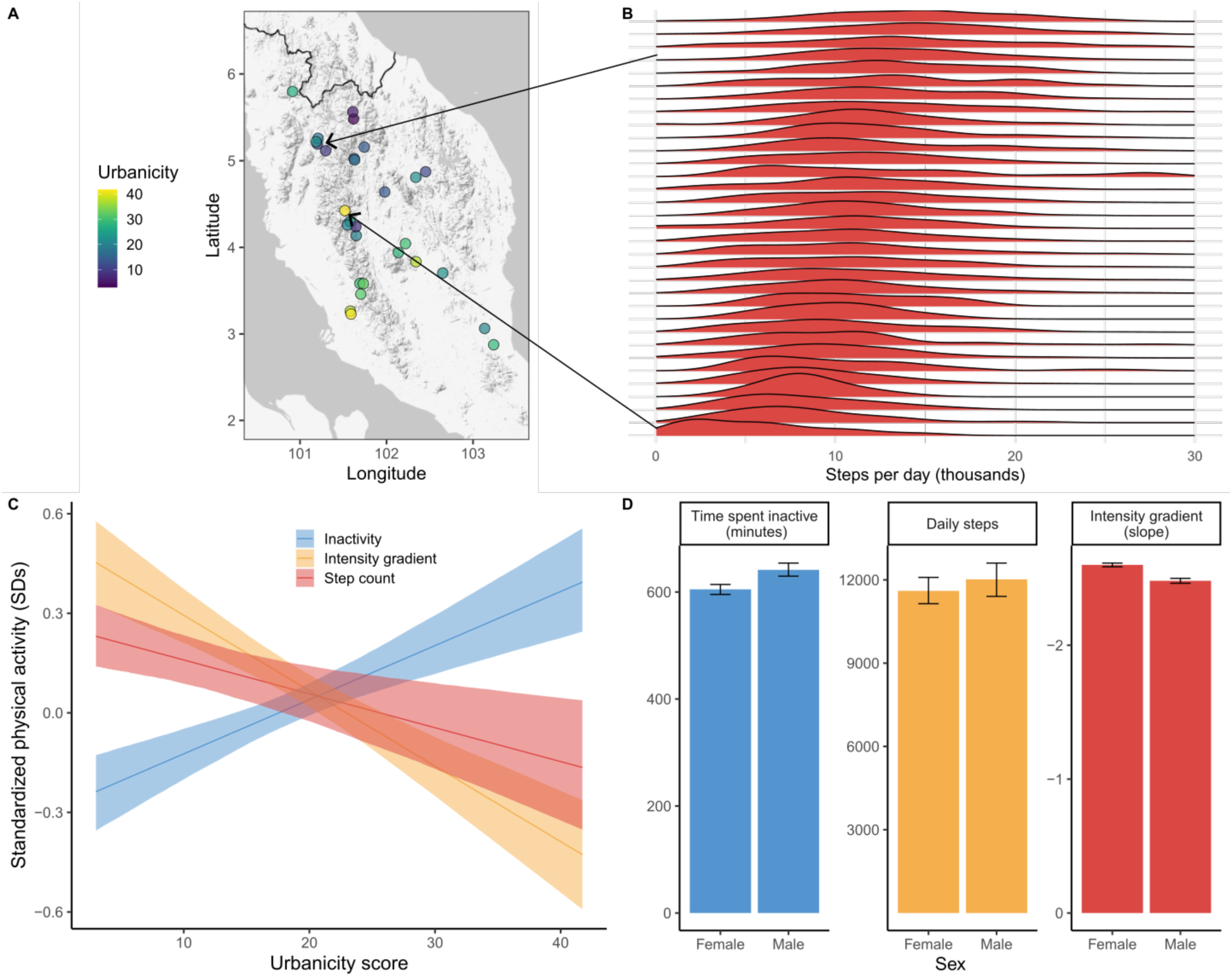
The effect of urbanicity on physical activity. (A) Map of Peninsular Malaysia showing the approximate locations of study villages, colored by the community-level urbanicity score (lighter and darker colors represent more urban and rural environments, respectively). (B) Density plots of physical activity (number of estimated steps per day) by village. Arrows show the location of two examples with very different levels of physical activity. (C) Predicted levels of physical activity (standardized) as a function of urbanicity. Lines and shading show mean predictions and 95% confidence intervals from multilevel model of physical activity outcomes as a function of age, sex, urbanicity, and a random intercept for individual (for female sex at average age of the sample). (D) Comparison of physical activity metrics between the sexes. Bars represent mean predicted response from posterior distributions ± 95% CIs. Note that the y-axis of the intensity gradient panel has been flipped such that more negative values are higher (a less negative value is indicative of more high intensity activity).

### Physical activity declines with urbanization are driven by less activity at later ages and reductions of low to moderate intensity activity

Physical activity tends to decrease with age across essentially all human populations, and the Orang Asli are no exception; steps per day, intensity gradient, and time spent in moderate or vigorous activity all decreased among older individuals, whereas time spent inactive or in light activity increased (Fig. 1B). To account for potential non-linear decreases in physical activity with age, we modeled age trends using a flexible spline function, yielding consistent results across the different physical activity outcomes. Average physical activity estimates remained relatively constant throughout adulthood, before beginning a rapid decline around age 50 (Fig. 1B).

Interestingly, however, the pace of decline after age 50 is heavily dependent on lifestyle variation. By age 75, fitted models show steep declines in physical activity to ∼3,400 steps/day (∼70.2% decline from peak) in the most urban locations, versus ∼12,300 steps/day (3.4% decline from peak) in the most traditional and least urban locations (Fig. 3A). Likewise, breakdowns of time spent in different intensities of physical activity reveal that urbanization promotes a shift from generally high levels of low to moderate intensity physical activity to greater time spent inactive (Fig. 3B; ꞵ [95% CI] for urbanicity effect from dirichlet regression relative to inactivity as baseline: ꞵvigorous = - 0.09 [-0.12, -0.05], ꞵmoderate = -0.05 [-0.09, -0.02], ꞵlight = -0.03 [-0.06, -0.01]). Vigorous physical activity is rare at both ends of the urbanization spectrum captured here. These findings indicate that the effects of urbanization on physical activity are age-dependent and predominantly driven by less time spent in low- to moderate-rather than high-intensity activities.

**Figure 3:**
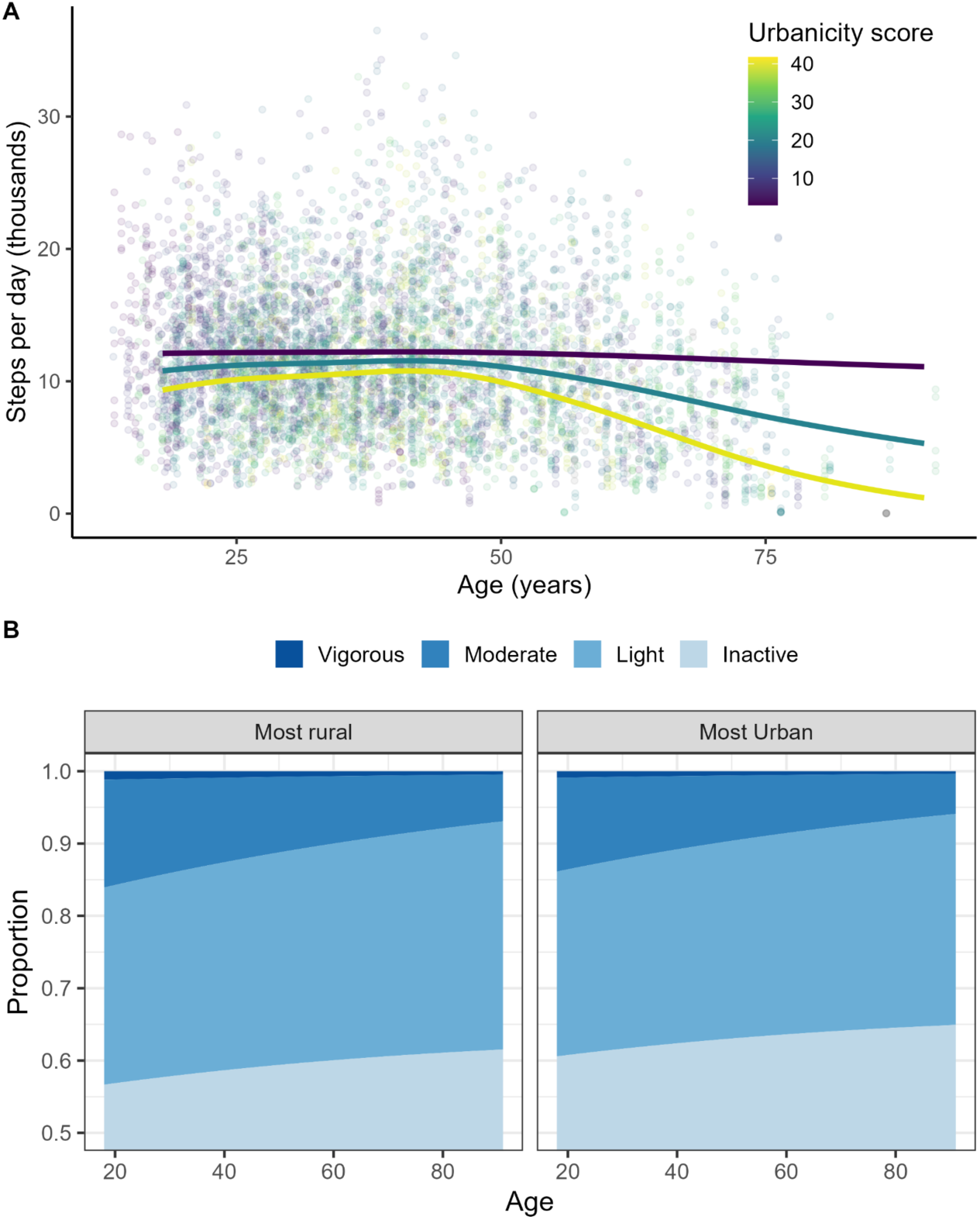
(A) Age-related decreases in physical activity begin around age 50 and occur more steeply in more urban environments. Each point is a person-day, colored by urbanicity score (yellow = more urban, purple = less urban), with lines representing model predictions from generalized additive mixed models of daily step counts as a function of an interaction between age (as a thin-plate spline) and urbanicity, adjusting for sex and a random intercept for repeat observations of individuals. Step counts were square-root transformed to improve normality and model fitting, with model predictions back-transformed for plotting. (B) The effects of urbanicity on time spent in different intensities of physical activity. Mean predicted proportions of time for individuals in the least and most urban environments, generated from a multilevel dirichlet regression with fixed effects of age, sex, and urbanicity score plus a random intercept for repeat observations of individuals. Note that the y-axis starts at 0.5 to bolster visualization of differences (all areas between 0-0.5 are inactivity).

### Physical activity declines and urbanization are associated with worse cardiometabolic health

Measurements of physical activity and lifestyle were paired with multiple cardiometabolic health variables for each participant, including measures of body composition (body weight, body fat percentage, waist circumference, waist to hip ratio, BMI, BRI, obese (y/n)), blood-based biomarkers (total cholesterol, LDL cholesterol, HDL cholesterol, triglycerides, prediabetes (y/n), diabetes (y/n)), and blood pressure (systolic, diastolic, hypertension (y/n)) (Materials and Methods). Health data were collected by a mobile team of professionally trained doctors and nurses during clinics that also provided free medical care to the entire community.

Consistent with our previous work (*22*), we found that cardiometabolic health among the Orang Asli was significantly worse in more urban environments. Specifically, weight, percent body fat, waist circumference, waist:hip ratio, BMI, and BRI, obesity, blood lipids (total cholesterol, LDL, triglycerides), systolic blood pressure, and rates of pre-diabetes and diabetes all increased significantly with increasing urbanicity score (Fig. 4A). We additionally found that metrics indicating greater levels of physical activity were associated with better cardiometabolic health across many, but not all, of the same outcome variables (Fig. 4B). For example, step counts, intensity gradient, and time spent inactive were all significant predictors in the expected direction for weight, percent body fat, waist circumference, BMI, BRI, triglycerides, diastolic blood pressure, and obesity. The remaining physical activity metrics demonstrated similar trends overall (Fig. S1), with some metrics, especially self-affinity of acceleration signals (DFA), generally having smaller magnitude effects for many outcomes (i.e., waist circumference). Further, multiple regression models simultaneously estimating the effects of inactivity and step counts or intensity gradient across biomarkers generally demonstrate independent, additive effects (Fig. S2).

**Figure 4:**
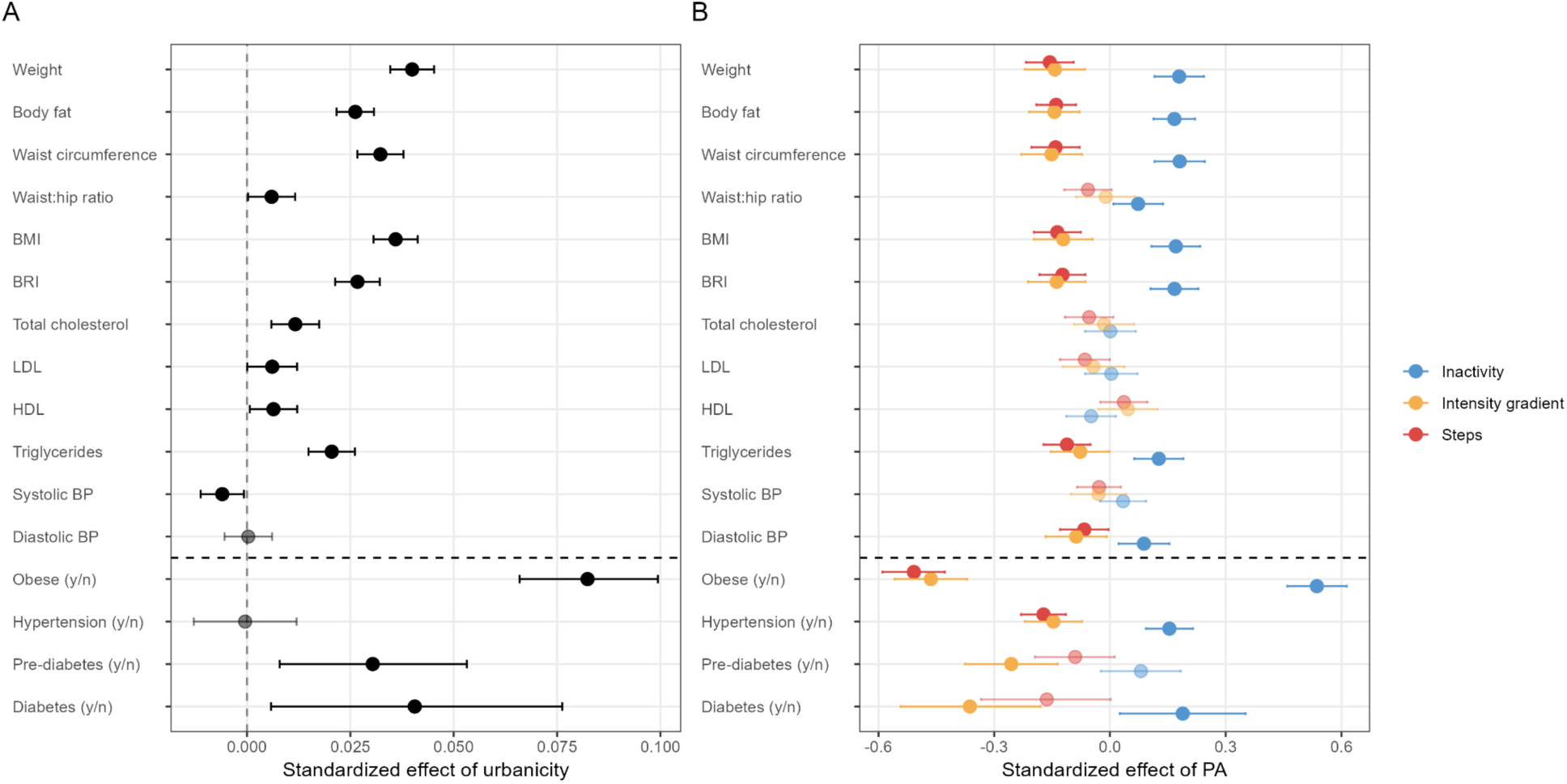
The effects of (A) urbanicity and (B) physical activity on cardiometabolic health outcomes. Points and error bars represent standardized effect size estimates (in units of standard deviations) and 95% confidence intervals from models of average physical activity metrics per day, adjusting for age*sex interaction. Estimates below the horizontal dashed line represent binary outcomes, which were not standardized for analysis. Transparency indicates non-significant effects (at P > 0.05) and colors in (B) show outcomes for four selected physical activity metrics.

### Physical activity as a mediator of urbanicity effects on health

In addition to analyzing the independent effects of both urbanicity and physical activity on cardiometabolic health, a central goal of our work was to determine whether physical activity was a mediator of the relationship between urbanicity and health (Fig. 5A). This is because many features of lifestyles that contribute to higher urbanicity scores, such as access to electricity and greater exposure to formal education, are unlikely to directly cause worse cardiometabolic health. Instead, urbanicity is considered to be a proxy for a suite of lifestyle variables that can influence health outcomes through multiple interconnected pathways, including potentially via physical activity patterns. To address this possibility, we employed formal mediation analyses with the goal of parsing out the extent to which urbanicity effects on health can be explained by associated changes to physical activity.

**Figure 5:**
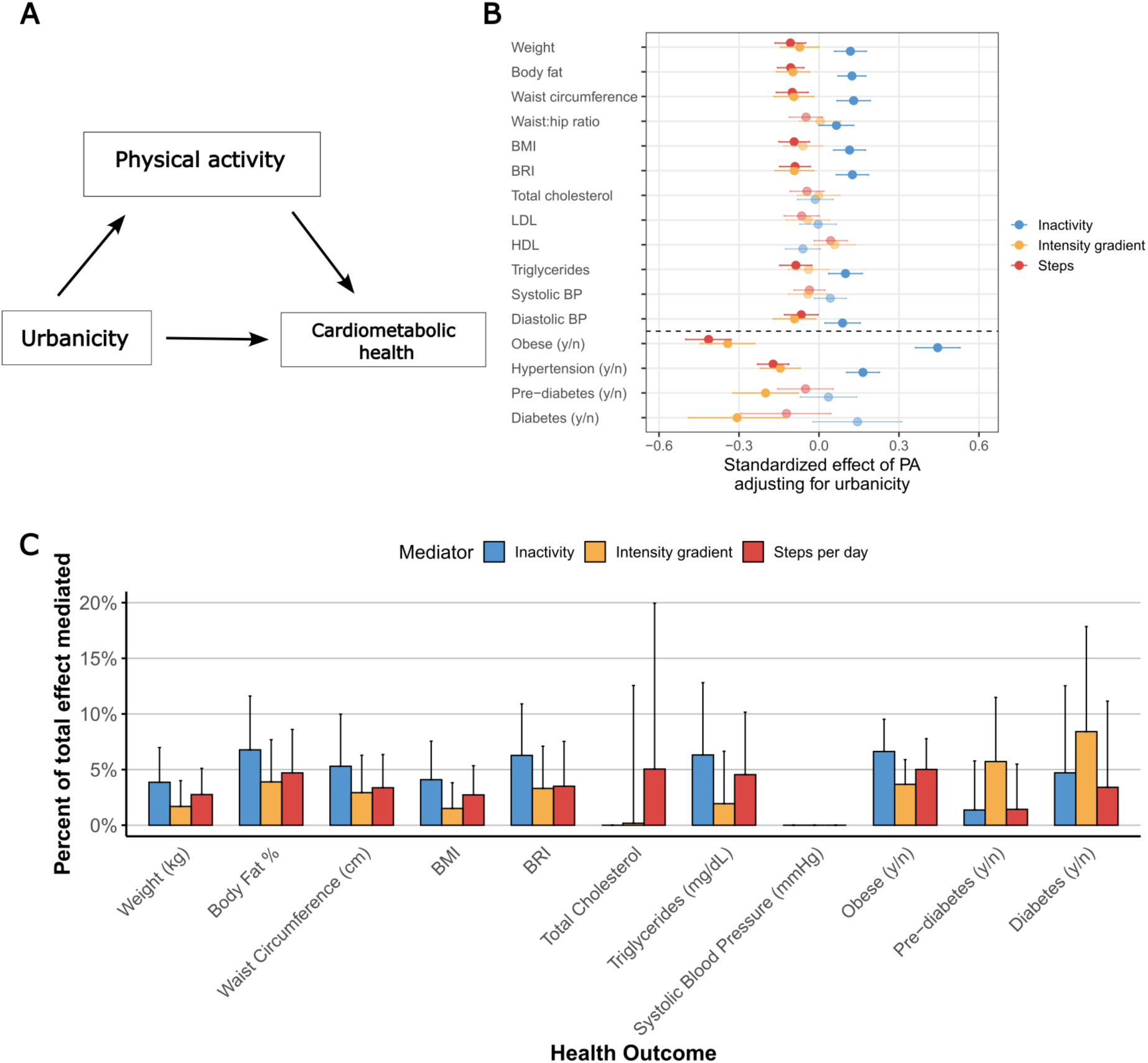
(A) Conceptual diagram for mediation analyses, showing physical activity as a potential mediator of the relationship between cardiometabolic health. (B) Forest plot of standardized coefficients ± 95% confidence intervals in regressions adjusting for the effect of urbanicity. (C) The percentage of the total urbanicity effect mediated by different physical activity metrics. Error bars represent upper 95% confidence intervals. Estimates of negative mediation (suppression) were set to zero for plotting.

Overall, physical activity was a significant mediator of urbanicity effects on most cardiometabolic health outcomes, but with generally small effect sizes (Fig. 5). Specifically, daily step counts were a significant mediator for 7 out of 11 outcomes, intensity gradient was a significant mediator for 6 out of 11 outcomes, and time spent inactive was a significant mediator for 7 out of 11 outcomes. For cases where physical activity was a significant mediator of urbanicity effects on health, the proportion of the total effect mediated was always small: across all cardiometabolic outcomes and physical activity mediators, the median proportion mediated was only 4.1% (range: 1.7 - 8.4%). These results show that the effects of urbanicity and physical activity on cardiometabolic health are largely independent (Fig. 5C). Physical inactivity was the strongest mediator overall across the different health outcomes, although only by small margins (and not for outcomes such as total cholesterol or pre-diabetes/diabetes).

## Discussion

Abundant comparative evidence suggests that humans evolved in the context of very high levels of physical activity (*11*), especially compared to our closest primate relatives (*17*). This evolved propensity is readily apparent among numerous studies of small-scale, non-industrial societies today that tend to engage in large amounts of physical activity, in addition to having excellent cardiometabolic health (*1*, *14*, *18*, *30*). Given that physical activity is a reliable predictor of improved health and mortality outcomes in industrialized contexts (*31*), it is often assumed that high levels of physical activity contribute substantially to these favorable cardiometabolic profiles (*2–6*, *8*, *9*). Yet, rigorous objective measurements of physical activity among non-industrial societies are rare, and as a result the direct role of decreasing physical activity in driving deteriorating cardiometabolic health during lifestyle transitions to more urban, industrialized contexts remains poorly understood. Moreover, some recent findings have called into question the extent to which high physical activity in non-industrial societies drives their good cardiometabolic health (*12–14*, *18*).

Using a large dataset of objectively-measured physical activity and a variety of cardiometabolic health outcomes across an extreme lifestyle gradient, this study addresses several fundamental questions: (1) How does physical activity change during the transition from non-industrial to more urban, industrial lifestyles?, (2) Are higher physical activity levels associated with better biomarkers of cardiometabolic health?, and (3) Is physical activity a key mediator of the relationship between more urban, industrial lifestyles and worse health?

### Physical activity and urbanization

The Orang Asli villages in this study were selected to represent a wide range of lifestyle variation spanning remote-living communities that are heavily reliant on foraging and subsistence farming to urban communities living in more “built” environments with heavy involvement in the market economy and access to modern infrastructure, technology, and formal education. We find clear evidence that objectively measured physical activity is reduced in more urban settings, regardless of the metric used to characterize activity (e.g., step counts, intensity gradient, time spent inactive) (Figs. 2,3). Physical activity reductions are underscored by increasing time spent inactive, and decreasing time spent in all intensities (low, moderate, and vigorous) of activity (Fig. 3B). These findings closely mirror global trends reported for industrializing (e.g., developing) low or middle income nations like Malaysia where urbanization has been consistently linked to decreases in physical activity (*32–38*).

Interestingly, among certain highly industrialized and post-industrial nations, evidence suggests that physical activity may sometimes be *lower* in rural than urban settings (*39*), mainly as a result of greater opportunities for recreational physical activity and/or less reliance on personal motorized vehicles in some urban cases (*38*). Thus, it is possible that, on a global level, the relationship between physical activity and urbanization is somewhat U-shaped, with Orang Asli representing only the lower portion of the urbanization spectrum (i.e., without highly sedentary desk jobs), albeit the end of the spectrum represented by the bulk of the global population and which is most relevant over human evolutionary history.

A likely possibility is that, as observed in China and the United States (*33*, *40*, *41*), a large component of the changes in Orang Asli physical activity following urbanization are explained by shifts in occupations or work-related activities. Subsistence economies invariably require large amounts of low- and moderate-intensity activities such as walking long distances, manual agricultural tasks like planting, weeding, and harvesting, and the ongoing need to process food, build/repair tools, and maintain houses (*17*), whereas wage labor can directly result in either higher or lower levels of activity. Although some common jobs for more market-integrated Orang Asli such as rubber tapping or work on palm oil plantations require repetitive and oftentimes hard physical labor, other occupations (e.g., truck driving, retail work) entail large amounts of sedentary time.

One of our most striking results is that age-related declines in physical activity vary widely across the lifestyle gradient. As observed almost universally around the world (*42–45*), physical activity decreases (and inactivity increases) with age among the Orang Asli (Fig. 1B). However, rates of decline, especially after age 50, are significantly steeper in individuals with more urbanized lifestyles (Fig. 3A). In fact, declines for Orang Asli in the most urban settings (∼22% per decade from age 45 to 75 in step counts) were even greater than those reported in the largest epidemiological study to date, in which Doherty et al. (*46*) estimated a 7.5% decline per decade of acceleration vector magnitude for people in the UK Biobank. Likewise, the modest declines observed in older Orang Asli with the most traditional, non-industrial lifestyles appeared similar to findings from other subsistence populations, including Hadza hunter-gatherers in Tanzania (females, but not males (*18*, *30*)) and Tsimane forager-horticulturalists in Bolivia (*14*).

### Physical activity and cardiometabolic health

Higher levels of physical activity and less time spent inactive were significantly correlated with many of the cardiometabolic health outcomes examined, especially traits related to body composition and obesity (Figs. 4,5). Few blood biomarker or blood pressure variables were strongly associated with physical activity, although step counts and time spent inactive were significant predictors of triglycerides and the probability that an individual was hypertensive was lower with higher daily step counts (Fig. 5B). These findings recapitulate a robust literature associating physical activity to cardiometabolic health (*11*, *19*, *47*, *48*), and are consistent with the idea that reductions in activity may play a major role in the rapid rise of chronic cardiometabolic diseases among Orang Asli and likely other groups undergoing lifestyle transition (*2*). Due to the cross-sectional and observational nature of our study, we are limited in the ability to infer causality or rule out the possibility that people with poor cardiometabolic health are less likely to be physically active due to loss of functional mobility (*49*), thus reversing the direction of causality. Nonetheless, repeated longitudinal studies (*50*, *51*) and randomized controlled trials (*48*) bolster our hypothesized causal pathway (Fig. 5A), which does not exclude the possibility that poor cardiometabolic health in the most extreme cases causes reductions in physical activity.

Stronger and more consistent associations between physical activity and body composition, versus blood pressure or lipids, are potentially a reflection of the relative roles of physical activity vis a vis other factors such as diet in determining these traits.This finding was unexpected, but not unprecedented. For example, Leon and Sanchez (*52*) aggregated published studies to demonstrate markedly inconsistent results between exercise/endurance training and blood lipids, whereas “all lipid parameters generally improved in studies in which there was a substantial weight loss (≥ 4 kg), usually associated with a concomitant hypocaloric diet.” Similarly, strong evidence links blood pressure to dietary factors (*53*), and in some cases with stronger effects than physical activity (*54*), though both are likely necessary components of hypertension control and prevention (*55*). Regardless, these findings raise the possibility that Orang Asli living more urbanized lifestyles may, at least in some cases, exhibit higher rates of obesity without experiencing a commensurate increase in other facets of metabolic syndrome, such as hypertension and hyperlipidemia.

Some researchers have suggested that, given our long evolutionary history of likely having engaged in universally high levels of physical activity compared to other primates, active lifestyles may be necessary for protecting health and functional ability, especially as we age (*11*, *56*). The strong associations observed here between objectively-measured physical activity and health, despite having an average of only ∼1 week of data per individual and the possibility of environmental changes occurring over the lifecourse (*57*), supports this general hypothesis. Our data further demonstrate that these associations persist even in quite rural and remote settings, where one might expect physical activity effects to have plateaued above generally high levels. Although this highlights critical health effects in humans, comparative (across species) tests remain necessary to determine whether evolution has more powerfully coupled physical activity to health due our history as “high-throughput” hunting and gathering species (*17*).

### Urbanization, physical activity, and cardiometabolic health

Our last major goal was to determine how much of the observed effects of urbanicity on cardiometabolic health (*22*) are mediated specifically by physical activity (Fig. 5A). The urbanicity index employed here does not directly generate worse cardiometabolic health; rather, its components (i.e., proportion of people with electricity in a village, housing type, levels of education) must correlate with lifestyle factors that are mechanistically linked to health outcomes. Previous studies demonstrating excellent cardiovascular health in subsistence populations commonly point to diet and physical activity as the key mediating variables (*8*), but these pathways are rarely investigated explicitly.

Formal mediation analyses paint a surprising picture; physical activity is a significant mediator for many health outcomes, but mediates only a small proportion (median: 4.1%) of the measured effect of urbanicity for nearly all relevant health outcomes (those that were significantly predicted by urbanicity; Fig. 5C). This suggests that physical activity only accounts for a minor portion of the variance in cardiometabolic health that correlates with urbanicity.

However, this is not because the effects of physical activity are merely subsumed by urbanicity. Although reduced in magnitude (Fig. 5B, Fig. S3), physical activity remains a robust, independent predictor of many cardiometabolic outcomes when controlling for urbanicity. We hypothesize that this result stems from the fact that the relationship between physical activity and health may be only weakly coupled with urbanization. For example, people in highly urban environments of low socioeconomic status may be likely to work jobs requiring exhausting physical activity, whereas some individuals in very rural environments may nonetheless gain ready access to labor-saving devices, such as chainsaws, motorbikes, or water filtration systems. As a result, it is clear that public health interventions should not overlook remote communities in which reduced physical activity may well appear in advance of obvious environmental changes.

## Conclusions

This study demonstrates that lifestyle changes with increasing urbanization are associated with both decreases in objectively-measured physical activity and concomitant worsening of cardiometabolic health. Despite physical activity being a common explanation for relationships between urbanization and health, physical activity appears to show independent effects and mediates only a small proportion of the total urbanization effects on most biomarkers in this study. In addition, age-related physical activity declines in older adults are notably more severe in more urban environments, suggesting that more subsistence-based lifestyles may be especially protective for cardiometabolic health of people above age 50 and raising the possibility of targeted interventions to raise physical activity among older adults in urban or peri-urban environments. Taken together, our findings support the general inference that recent global reductions in physical activity represent an evolutionarily novel challenge for our species, and that high levels of physical activity are an important contributor to the generally high quality of cardiometabolic health that has long been observed among subsistence populations around the world (*1*).

The proliferation of studies systematically collecting objective measures of physical activity, particularly those garnering acceleration data that can be subjected to standardized analyses, offer tremendous promise for the future of research on physical activity and health. In particular, there is much to be learned from studies of diverse populations experiencing disparate environmental conditions, especially as the expansion of globalization continues to generate “natural” experiments that can be leveraged to explore variation in the effects of changing physical activity profiles. In addition to large-scale synthesis of data and findings from small-scale subsistence societies to inform our understanding of what kinds of physical activity levels are representative for our species over evolutionary time, ripe areas of future study include longitudinal research in groups like the Orang Asli experiencing rapid lifestyle change, circadian analyses explicitly investigating the times of day when physical activity reductions are most acute in urban environments (*46*, *58*), and investigation of the relative impacts of light/moderate/vigorous activities on health in subsistence contexts where recreational physical activity is low. Such research has only become more consequential following broad theoretical and empirical developments challenging traditional models of the relationship between physical activity and total daily energy expenditure (*59*), and will help clarify the particular ways in which modern lifestyles are contributing to the global rise of chronic non-communicable disease.

## Materials and Methods

### Study Population

The Orang Asli, or “original people,” are comprised of approximately 19 distinct ethnolinguistic groups that collectively represent the Indigenous peoples of Peninsular Malaysia. Data for this study come from 34 villages that are predominantly occupied by the following groups: Batek, Batek Nogn, Jahai, Jahut, Jakun, Kensiu, Semai, Semaq Beri, Temiar, and Temuan. These groups have long been separated by anthropologists into three major categories based primarily on language, subsistence type, and phenotypic differences (*60*, *61*).

The Batek, Batek Nong, Jahai, and Kensiu are generally categorized by anthropologists as “negrito” or “Semang” (both terminologies with controversial colonial histories), speak languages in the Northern Aslian sub-branch of the Austroasiatic language family, and traditionally lived as mobile nomadic hunter-gatherers and traders of forest products primarily in the north and central regions of Peninsular Malaysia (*62*). The Semai, Semaq Beri, and Temiar are collectively referred to as “Senoi”, speak languages in the Central Aslian subbranch of the Austroasiatic language family, live mainly in the central regions of Malaysia including highland areas of the main range, and traditionally lived sedentary or semi-nomadic lifestyles practicing swidden horticulture (mixed crops with staples primarily consisting of millet, cassava, sweet potato, hill rice, and banana) supplemented with fishing and hunting. In contrast, the Jakun and Temuan live primarily in the south and are categorized as “Proto-Malay”, speak Austronesian languages, are phenotypically more similar to predominant ethnic Malay populations, and practice mixed subsistence strategies including permanent agriculture, riverine or coastal fishing, and wage labor or professional jobs. The Proto-Malay are generally more market integrated compared to other Orang Asli. In addition, all of these Orang Asli groups have rich and varied histories that have resulted in distinct religions, coresidence patterns, marriage practices, mobility patterns, and other important aspects of socioecology (for more information on these groups, we refer readers to: (*60–68*)).

A notable feature of the Orang Asli environment is the dramatic lifestyle gradient that exists today, ranging from semi-nomadic practitioners of foraging lifestyles to fully sedentarized groups living in urban or peri-urban centers who are fully engaged with the national market economy (*23*). Among the most urban groups, Orang Asli tend to be of relatively low socioeconomic status, with low levels of education compared to majority ethnic groups and low rates of higher paying, sedentary office jobs. Although all Orang Asli today have some engagement with the market economy, the nature and extent of this engagement is highly variable. For example, the economies of some Temuan communities in close proximity to Kuala Lumpur are highly dependent on wage labor, with community members working a variety of wage labor jobs (i.e., shopkeeper, hotel worker, resort staff, plantation worker), whereas some Batek and Jahai communities have only a minority of individuals working wage labor jobs (in sectors such as tourism or tapping rubber), with others instead relying on a mix of subsistence foraging, small-scale cropping, and the collection and trade of forest products. The lifestyle gradient that exists today is largely a result of Malaysia’s recent history of rapid economic development; prior to the 1970’s and 1980’s, the interior regions of Peninsular Malaysia remained relatively inaccessible with little built infrastructure (with notable exceptions in areas developed for mining operations, mainly gold and tin). After this time, however, major development projects began aimed at increasing exploitation of timber and mining, in addition to the expansion of large-scale oil palm and rubber agriculture. Combined with substantial oil revenues, in the past 50 years Malaysia has thus been propelled through an extremely rapid transition toward developed status. Orang Asli have remained relatively marginalized throughout this process with a high proportion currently living in poverty, but the extent of economic development is highly variable across communities and is heavily dependent on geographical proximity to major population centers.

### Study design

This study is part of the *Orang Asli Health and Lifeways Project (OAHeLP)*, a long-term study of behavior, culture, environment, and health among Orang Asli throughout Peninsular Malaysia (*23*). Data are collected from consenting adults during mobile health clinics which travel to study communities to provide free healthcare onsite. In addition to providing treatment, a team of doctors, assistants, and anthropologists conduct lifestyle interviews and comprehensive health screenings, including analysis of blood-based biomarkers from venous blood draw using point-of-care devices. 16 cardiometabolic phenotypes were measured or derived for all participants. These include weight (kg), body fat percentage (from bioimpedance), waist circumference (cm), body mass index (BMI), body roundness index (BRI) (*69*), waist-to-hip ratio, total cholesterol, high-density and low-density lipoprotein (HDL, LDL) cholesterol, triglycerides (mg/dL), and systolic and diastolic blood pressure (mmHg). We also characterized whether the individual fit criteria for being hypertensive (systolic blood pressure ≥ 130 or diastolic blood pressure ≥ 80), obese (BMI ≥ 30), prediabetic (6.5 > HbA1c ≥ 5.7 or 125 > non-fasting glucose ≥ 199 in few cases where HbA1c not available), or diabetic (HbA1c ≥ 6.5 or non-fasting glucose ≥ 200 in few cases where HbA1c not available). “Urbanicity” scores were derived from interviews as described in Watowich et al. (*22*) and Table S1. All data in this study were collected between 2020 and 2024. Detailed data collection protocols are described previously (*23*) (Wallace et al., 2022).

### Accelerometry

At the end of research clinics, all participants were invited to wear research grade accelerometers (Axivity AX3) on the non-dominant wrist for a period of one week, after which time the devices were collected and returned to researchers. Accelerometry is restricted to periods outside of research clinics to ensure that behavior is not altered by the presence of our research team. Accelerometers are configured to record information at a sampling frequency of 100 Hz with a dynamic range of 8 *g*.

### Accelerometry Data Processing

Raw accelerometry files were downloaded and pre-processed to remove any days recorded before or after participants wore the devices. Next, we used the *GGIR* package (v3.2.6) (*24*, *70*) to process the data into 5-second epochs for analysis using default auto-calibration and imputation algorithms. Non-wear time was also estimated using built-in *GGIR* algorithms, and we defined a valid day for inclusion as those in which ≥ 16 hours of wear time were detected. A *GGIR* configuration file describing the full set of parameters used in our analysis is provided as supplementary material.

We extracted seven summary metrics in order to capture distinct aspects of physical activity (Table 1). These include two metrics for overall PA (ENMO and daily step counts), five metrics capturing the intensity of activity (time spent inactive or in light, moderate, or vigorous activity, and intensity gradient), and a complexity-based measure of self-similarity characterizing patterns of physical activity over time that has been previously demonstrated to strongly predict mortality (*71*). Physical activity metrics were calculated at each person-day (with days calculated from midnight - midnight), but for some analyses averages were taken at the individual level.

### Statistical Analyses

The effects of age on physical activity outcomes were estimated using generalized additive mixed models with age terms modeled using thin-plate splines and random intercepts for repeat observations of individuals, using data in which each row represented a person-day for a given physical activity metric. To assess the effect of urbanicity on physical activity, we fit Bayesian multilevel models of physical activity outcomes as a function of age (thin-plate spline), sex, urbanicity, and the interaction of age and urbanicity with a random intercept for individual (Figs. 2,3). Models were fit using the *brms* package in R using weakly informative priors. Standard diagnostics indicated no problems during estimation (R^ ≅ 1, good mixing, and sufficient effective sample sizes).

We analyzed the effect of physical activity on cardiometabolic health outcomes at the person-level by first collapsing physical activity metrics across days into the average value, and standardizing (subtracting the mean and dividing by the standard deviation) continuous variables for comparison. Prior to standardization, several variables with highly skewed distributions were transformed to approximate normality: square-root transformations were applied to total cholesterol, LDL, HDL, and triglycerides; systolic blood pressure was natural log transformed. Variables with binary outcomes (e.g., hypertension) were not transformed or standardized. We then ran a series of linear models regressing the cardiometabolic outcomes on either urbanicity (Fig. 4A) or physical activity metrics (Fig. 4B), separately, adjusting for an age*sex interaction. Sample sizes are provided in Table S2.

Lastly, we conducted formal mediation analysis to examine how empirical estimates of physical activity mediate the relationship between urbanicity and cardiometabolic health using the R package *mediation* (Fig. 5). Specifically, this involves fitting multiple models, one estimating physical activity as a function of urbanicity and another with cardiometabolic health outcomes as a function of both physical activity and urbanicity scores (with all models including additional age*sex interaction covariates). Model parameters are then compared to estimate the average causal mediation effect and the proportion of the total urbanicity effect mediated specifically by a given physical activity metric. A bootstrapping procedure was used to determine significance of the mediation. Mediation tests were only performed if prior analyses indicated a significant relationship (p<0.05) between urbanicity and a given cardiometabolic outcome.

All analyses were conducted in the R statistical programming environment version 4.5.0 (*72*). All model outputs are provided in Tables S3-5.

### Ethical Approvals

This research was approved by the Medical Review and Ethics Committee of the Malaysian Ministry of Health (protocol ID: NMRR-20-2214-55565), the Malaysian Department of Orang Asli Development (permit ID: JAKOA.PP.30.052 JLD 21 (98)), and the Institutional Review Board of Vanderbilt University (protocol ID: 212175). Informed written consent was provided by all individuals at the time of data collection.

## Data Availability Statement

OA HeLP’s highest priority is the minimization of risk to study participants. OA HeLP adheres to the ‘CARE Principles for Indigenous Data Governance’ (Collective Benefit, Authority to Control, Responsibility, and Ethics). OA HeLP is also committed to the ‘FAIR Guiding Principles for scientific data management and stewardship’ (Findable, Accessible, Interoperable, Reusable). To adhere to these principles while minimizing risks, individual-level data are stored in the OA HeLP protected data repository, and are available through restricted access. Requests for de-identified, individual-level data should take the form of an application that details the exact uses of the data and the research questions to be addressed, procedures that will be employed for data security and individual privacy, potential benefits to the study communities and procedures for assessing and minimizing stigmatizing interpretations of the research results. Requests for de-identified, individual-level data will require institutional IRB approval (even if exempt). OA HeLP is committed to open science and the project leadership is available to assist interested investigators in preparing data access requests (see orangaslihealth.org for further details and contact information).

## Acknowledgements

We thank the Orang Asli participants who have generously allowed us to work in their communities and have shared tremendous amounts of knowledge over the years. We also thank the Universiti Malaya Department of Parasitology for hosting our research team, the Centre for Malaysian Indigenous Studies (CMIS) at Universiti Malaya for critical logistical support, and Jordan Hoffman for help with data entry. This work was supported by the National Science Foundation (BCS-2142090), the Canadian Institute for Advanced Research (Azrieli Global Scholars Program), and the Pew Charitable Trusts (Pew Biomedical Scholars Program). The funders had no role in study design, data collection and analysis, decision to publish, or preparation of the manuscript.

