## Supplemental Figures and Tables for "Physical activity and cardiometabolic health across an extreme lifestyle gradient"

### Supplementary Information for Kraft et al. “Physical activity and cardiometabolic health across an extreme lifestyle gradient”

**Table of contents:**


- Figure S1. Main analysis results including all metrics of physical activity
- Figure S2. The effects of daily step counts and intensity gradient on cardiometabolic health outcomes controlling for inactivity
- Figure S3. Adjusting for urbanicity minimally changes the relationship between physical activity and cardiometabolic health
- Table S1. Information on urbanicity score calculations
- Table S2. Sample sizes for comparisons between physical activity and cardiometabolic health outcomes
- Table S3. Full model results for analyses describing urbanicity effects on physical activity (provided as an HTML file)
- Table S4. Full model results for analyses describing physical activity effects on health (provided as an HTML file)
- Table S5. Full model results for analyses describing physical activity effects on health, controlling for urbanicity (provided as an HTML file)

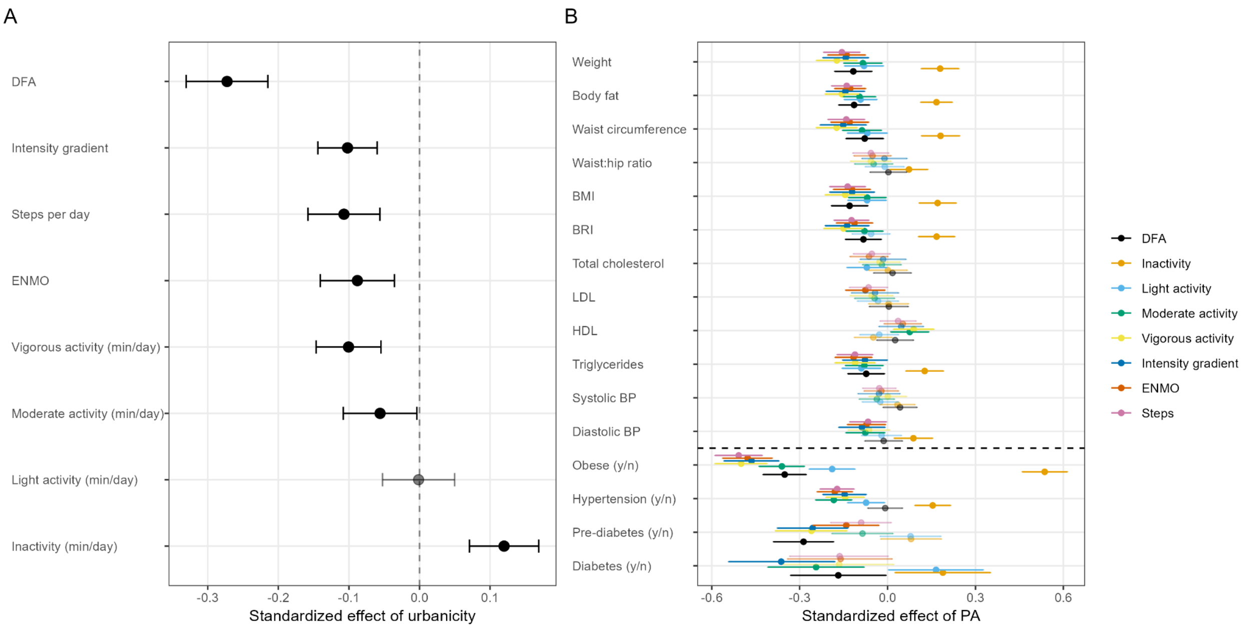


**Figure S1. Main analysis results including all metrics of physical activity.** (A) The standardized effect of urbanicity on different physical activity metrics, adjusting for age x sex interactions with a random intercept to account for repeat observations of individuals. Bars represent 95% confidence intervals. (B) Forest plot of standardized effects of all physical activity variables measured in this study on the full suite of cardiometabolic health outcomes. All models adjust for age x sex interactions (as in Fig. 4 in the main text).

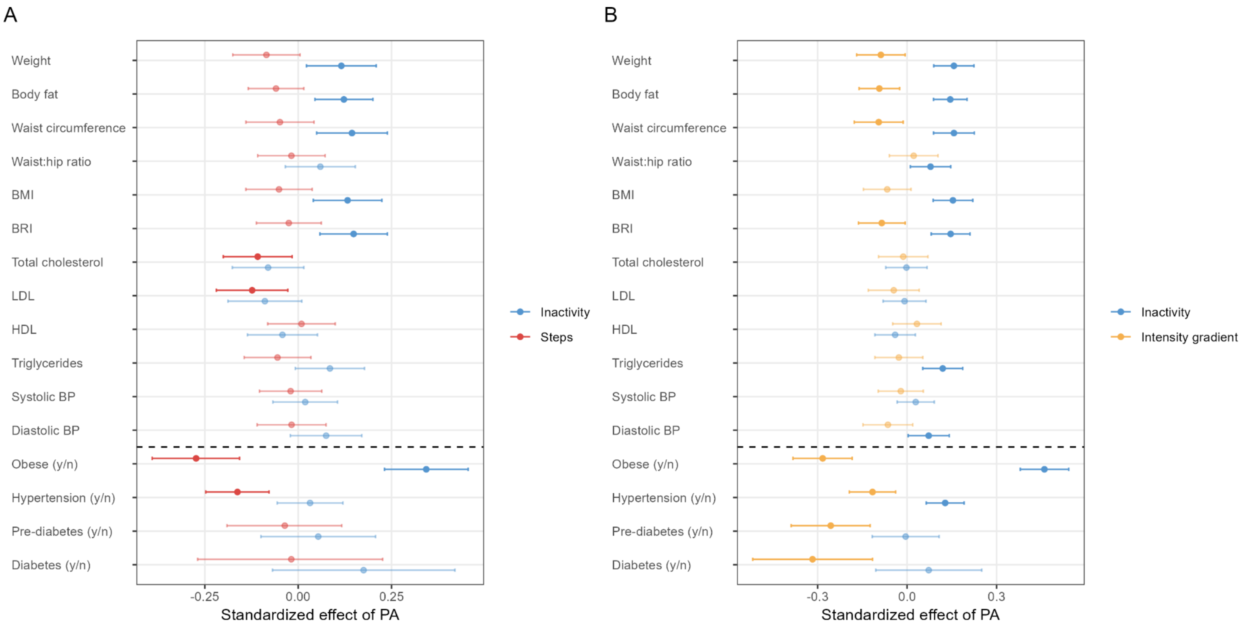


**Figure S2. The effects of (A) daily step counts and (B) intensity gradient on cardiometabolic health outcomes controlling for inactivity**. Forest plots represent model estimated effects (plus confidence intervals) from multiple regression models that included the focal physical activity predictor as well as time spent inactive. All models are weighted by the number of valid measurement days of physical activity and adjusted for an age x sex interaction.

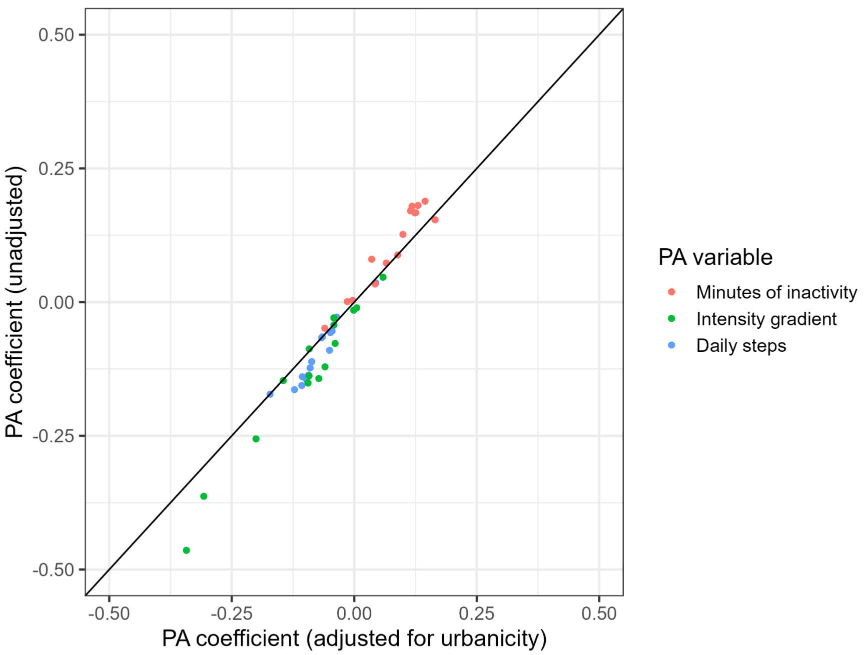


**Figure S3. Adjusting for urbanicity minimally changes the relationship between physical activity and cardiometabolic health.** The x-axis represents model coefficients for effects of physical activity (PA) variables on health outcomes in models adjusting for urbanicity score, whereas the y-axis represents coefficients from the same models but without adjusting for urbanicity score.

**Table S1. Information on urbanicity score calculations**

<u>These items were tallied to create an urbanicity score for each location:</u>

- Scaled population density
- 10 - (10*Proportion listed as not foraging, agriculture, fishing)
- 5*Proportion of households with flush toilets
- 5*Proportion of households with electricity
- 5*Proportion of households with television
- 5*Proportion of households with smart phones
- 10*Proportion of >40yr olds with some education
- 10*Proportion of <40yr olds with some education

<u>Population density was scaled as follows:</u>

| Score | Density per square kilometer |
|---|---|
| 0.67 | 0-99 |
| 1.33 | 100-199 |
| 2.00 | 200-299 |
| 2.67 | 300-399 |
| 3.33 | 400-499 |
| 4.00 | 500-999 |
| 4.67 | 1000-1999 |
| 5.33 | 2000-2999 |
| 6.00 | 3000-3999 |
| 6.67 | 4000-5999 |
| 7.33 | 6000-7999 |
| 8.00 | 8000-9999 |
| 8.67 | 10000-14999 |
| 9.33 | 15000-19999 |
| 10.00 | 20000-Inf |

#### Table S2. Sample sizes for comparisons between physical activity and cardiometabolic health outcomes

| Outcome | Predictor | Individuals |
|---|---|---|
| Body weight | Inactivity | 1048 |
| Body fat % | Inactivity | 1025 |
| Waist circumference | Inactivity | 1027 |
| Waist to hip ratio | Inactivity | 1022 |
| BMI | Inactivity | 1048 |
| BRI | Inactivity | 1027 |
| Total cholesterol | Inactivity | 1030 |
| LDL cholesterol | Inactivity | 964 |
| HDL cholesterol | Inactivity | 1030 |
| Triglycerides | Inactivity | 1030 |
| Systolic blood pressure | Inactivity | 1026 |
| Diastolic blood pressure | Inactivity | 1026 |
| Obese (y/n) | Inactivity | 1048 |
| Hypertension (y/n) | Inactivity | 1026 |
| Prediabetes (y/n) | Inactivity | 1022 |
| Diabetes (y/n) | Inactivity | 1022 |
| Body weight | Steps | 1053 |
| Body fat % | Steps | 1029 |
| Waist circumference | Steps | 1032 |
| Waist to hip ratio | Steps | 1027 |
| BMI | Steps | 1053 |
| BRI | Steps | 1032 |
| Total cholesterol | Steps | 1035 |
| LDL cholesterol | Steps | 970 |
| HDL cholesterol | Steps | 1035 |
| Triglycerides | Steps | 1035 |
| Systolic blood pressure | Steps | 1031 |

| | | |
|---|---|---|
| Diastolic blood pressure | Steps | 1031 |
| Obese (y/n) | Steps | 1053 |
| Hypertension (y/n) | Steps | 1031 |
| Prediabetes (y/n) | Steps | 1026 |
| Diabetes (y/n) | Steps | 1026 |
| Body weight | Intensity gradient | 1053 |
| Body fat % | Intensity gradient | 1029 |
| Waist circumference | Intensity gradient | 1032 |
| Waist to hip ratio | Intensity gradient | 1027 |
| BMI | Intensity gradient | 1053 |
| BRI | Intensity gradient | 1032 |
| Total cholesterol | Intensity gradient | 1035 |
| LDL cholesterol | Intensity gradient | 970 |
| HDL cholesterol | Intensity gradient | 1035 |
| Triglycerides | Intensity gradient | 1035 |
| Systolic blood pressure | Intensity gradient | 1031 |
| Diastolic blood pressure | Intensity gradient | 1031 |
| Obese (y/n) | Intensity gradient | 1053 |
| Hypertension (y/n) | Intensity gradient | 1031 |
| Prediabetes (y/n) | Intensity gradient | 1026 |
| Diabetes (y/n) | Intensity gradient | 1026 |
