## Supplementary Table 3 for "Physical activity and cardiometabolic health across an extreme lifestyle gradient"

|  | Weight | | | | Body fat | | | | Waist circumference | | | | Waist:hip ratio | | | | BMI | | | | BRI | | | | Total cholesterol | | | | LDL | | | | HDL | | | | Triglycerides | | | | Systolic BP | | | | Diastolic BP | | | | Obese (y/n) | | | | Hypertension (y/n) | | | | Pre-diabetes (y/n) | | | | Diabetes (y/n) | | | |
| --- | --- | --- | --- | --- | --- | --- | --- | --- | --- | --- | --- | --- | --- | --- | --- | --- | --- | --- | --- | --- | --- | --- | --- | --- | --- | --- | --- | --- | --- | --- | --- | --- | --- | --- | --- | --- | --- | --- | --- | --- | --- | --- | --- | --- | --- | --- | --- | --- | --- | --- | --- | --- | --- | --- | --- | --- | --- | --- | --- | --- | --- | --- | --- | --- |
| Predictors | Estimates | std. Error | CI | p | Estimates | std. Error | CI | p | Estimates | std. Error | CI | p | Estimates | std. Error | CI | p | Estimates | std. Error | CI | p | Estimates | std. Error | CI | p | Estimates | std. Error | CI | p | Estimates | std. Error | CI | p | Estimates | std. Error | CI | p | Estimates | std. Error | CI | p | Estimates | std. Error | CI | p | Estimates | std. Error | CI | p | Odds Ratios | std. Error | CI | p | Odds Ratios | std. Error | CI | p | Odds Ratios | std. Error | CI | p | Odds Ratios | std. Error | CI | p |
| Intercept | -0.35 | 0.11 | -0.55 – -0.14 | **0.001** | 0.02 | 0.09 | -0.16 – 0.19 | 0.866 | -0.69 | 0.11 | -0.91 – -0.47 | **<0.001** | -0.81 | 0.11 | -1.03 – -0.58 | **<0.001** | -0.27 | 0.11 | -0.48 – -0.06 | **0.011** | -0.67 | 0.11 | -0.89 – -0.46 | **<0.001** | -0.73 | 0.12 | -0.96 – -0.51 | **<0.001** | -0.48 | 0.12 | -0.72 – -0.24 | **<0.001** | -0.22 | 0.12 | -0.45 – 0.00 | 0.054 | -1.00 | 0.11 | -1.22 – -0.77 | **<0.001** | -1.33 | 0.11 | -1.53 – -1.12 | **<0.001** | -0.76 | 0.12 | -0.99 – -0.53 | **<0.001** | 0.11 | 0.03 | 0.06 – 0.20 | **<0.001** | 0.17 | 0.05 | 0.10 – 0.29 | **<0.001** | 0.01 | 0.01 | 0.00 – 0.03 | **<0.001** | 0.00 | 0.00 | 0.00 – 0.01 | **<0.001** |
| Age | -0.01 | 0.00 | -0.02 – -0.01 | **<0.001** | -0.00 | 0.00 | -0.01 – 0.00 | 0.070 | 0.00 | 0.00 | -0.00 – 0.01 | 0.440 | 0.02 | 0.00 | 0.01 – 0.02 | **<0.001** | -0.01 | 0.00 | -0.01 – -0.00 | **<0.001** | 0.01 | 0.00 | 0.00 – 0.01 | **0.001** | 0.02 | 0.00 | 0.01 – 0.02 | **<0.001** | 0.01 | 0.00 | 0.01 – 0.02 | **<0.001** | 0.01 | 0.00 | 0.00 – 0.01 | **0.014** | 0.01 | 0.00 | 0.01 – 0.02 | **<0.001** | 0.03 | 0.00 | 0.03 – 0.04 | **<0.001** | 0.02 | 0.00 | 0.01 – 0.02 | **<0.001** | 0.98 | 0.01 | 0.96 – 0.99 | **0.002** | 1.05 | 0.01 | 1.03 – 1.06 | **<0.001** | 1.03 | 0.01 | 1.02 – 1.05 | **<0.001** | 1.04 | 0.01 | 1.01 – 1.06 | **0.010** |
| Sex | 0.18 | 0.16 | -0.14 – 0.50 | 0.267 | -1.48 | 0.14 | -1.75 – -1.20 | **<0.001** | -0.34 | 0.17 | -0.67 – -0.00 | **0.049** | 0.22 | 0.18 | -0.13 – 0.56 | 0.219 | -0.48 | 0.16 | -0.80 – -0.15 | **0.004** | -0.67 | 0.17 | -1.00 – -0.35 | **<0.001** | 0.13 | 0.18 | -0.22 – 0.48 | 0.483 | -0.16 | 0.19 | -0.53 – 0.20 | 0.385 | -0.75 | 0.18 | -1.10 – -0.41 | **<0.001** | 0.98 | 0.17 | 0.64 – 1.32 | **<0.001** | 1.02 | 0.16 | 0.70 – 1.34 | **<0.001** | 0.27 | 0.18 | -0.08 – 0.62 | 0.134 | 0.25 | 0.15 | 0.08 – 0.78 | **0.019** | 2.39 | 0.96 | 1.09 – 5.23 | **0.030** | 0.69 | 0.53 | 0.15 – 2.99 | 0.633 | 3.13 | 3.44 | 0.35 – 26.37 | 0.298 |
| Urbanicity score | 0.04 | 0.00 | 0.03 – 0.05 | **<0.001** | 0.03 | 0.00 | 0.02 – 0.03 | **<0.001** | 0.03 | 0.00 | 0.03 – 0.04 | **<0.001** | 0.01 | 0.00 | 0.00 – 0.01 | **0.040** | 0.04 | 0.00 | 0.03 – 0.04 | **<0.001** | 0.03 | 0.00 | 0.02 – 0.03 | **<0.001** | 0.01 | 0.00 | 0.01 – 0.02 | **<0.001** | 0.01 | 0.00 | 0.00 – 0.01 | **0.047** | 0.01 | 0.00 | 0.00 – 0.01 | **0.029** | 0.02 | 0.00 | 0.01 – 0.03 | **<0.001** | -0.01 | 0.00 | -0.01 – -0.00 | **0.025** | 0.00 | 0.00 | -0.01 – 0.01 | 0.915 | 1.09 | 0.01 | 1.07 – 1.10 | **<0.001** | 1.00 | 0.01 | 0.99 – 1.01 | 0.944 | 1.03 | 0.01 | 1.01 – 1.05 | **0.008** | 1.04 | 0.02 | 1.01 – 1.08 | **0.023** |
| Age:Sex | 0.00 | 0.00 | -0.01 – 0.01 | 0.674 | 0.01 | 0.00 | 0.00 – 0.01 | **0.015** | 0.00 | 0.00 | -0.00 – 0.01 | 0.341 | 0.00 | 0.00 | -0.01 – 0.01 | 0.801 | 0.00 | 0.00 | -0.00 – 0.01 | 0.264 | 0.00 | 0.00 | -0.01 – 0.01 | 0.581 | -0.01 | 0.00 | -0.02 – -0.00 | **0.021** | -0.00 | 0.00 | -0.01 – 0.00 | 0.296 | 0.01 | 0.00 | -0.00 – 0.02 | 0.091 | -0.01 | 0.00 | -0.02 – -0.00 | **0.002** | -0.02 | 0.00 | -0.02 – -0.01 | **<0.001** | -0.00 | 0.00 | -0.01 – 0.00 | 0.253 | 1.01 | 0.01 | 0.98 – 1.04 | 0.411 | 0.99 | 0.01 | 0.97 – 1.01 | 0.185 | 1.00 | 0.02 | 0.97 – 1.03 | 0.931 | 0.98 | 0.02 | 0.93 – 1.02 | 0.276 |
| Observations | 1124 | | | | 1105 | | | | 1091 | | | | 1086 | | | | 1124 | | | | 1091 | | | | 1094 | | | | 1026 | | | | 1094 | | | | 1094 | | | | 1090 | | | | 1090 | | | | 1124 | | | | 1091 | | | | 1086 | | | | 1086 | | | |
