## Supplementary Table 4 for "Physical activity and cardiometabolic health across an extreme lifestyle gradient"

|  | Weight | | | | Body fat | | | | Waist circumference | | | | Waist:hip ratio | | | | BMI | | | | BRI | | | | Total cholesterol | | | | LDL | | | | HDL | | | | Triglycerides | | | | Systolic BP | | | | Diastolic BP | | | | Obese (y/n) | | | | Hypertension (y/n) | | | | Pre-diabetes (y/n) | | | | Diabetes (y/n) | | | |
| --- | --- | --- | --- | --- | --- | --- | --- | --- | --- | --- | --- | --- | --- | --- | --- | --- | --- | --- | --- | --- | --- | --- | --- | --- | --- | --- | --- | --- | --- | --- | --- | --- | --- | --- | --- | --- | --- | --- | --- | --- | --- | --- | --- | --- | --- | --- | --- | --- | --- | --- | --- | --- | --- | --- | --- | --- | --- | --- | --- | --- | --- | --- | --- | --- |
| Predictors | Estimates | std. Error | CI | p | Estimates | std. Error | CI | p | Estimates | std. Error | CI | p | Estimates | std. Error | CI | p | Estimates | std. Error | CI | p | Estimates | std. Error | CI | p | Estimates | std. Error | CI | p | Estimates | std. Error | CI | p | Estimates | std. Error | CI | p | Estimates | std. Error | CI | p | Estimates | std. Error | CI | p | Estimates | std. Error | CI | p | Odds Ratios | std. Error | CI | p | Odds Ratios | std. Error | CI | p | Odds Ratios | std. Error | CI | p | Odds Ratios | std. Error | CI | p |
| Intercept | 0.42 | 0.11 | 0.21 – 0.63 | **<0.001** | 0.55 | 0.09 | 0.38 – 0.73 | **<0.001** | -0.08 | 0.11 | -0.29 – 0.14 | 0.482 | -0.66 | 0.11 | -0.88 – -0.45 | **<0.001** | 0.46 | 0.11 | 0.25 – 0.67 | **<0.001** | -0.14 | 0.10 | -0.34 – 0.07 | 0.189 | -0.50 | 0.11 | -0.72 – -0.28 | **<0.001** | -0.32 | 0.12 | -0.54 – -0.09 | **0.006** | -0.19 | 0.11 | -0.40 – 0.02 | 0.074 | -0.57 | 0.11 | -0.78 – -0.36 | **<0.001** | -1.41 | 0.10 | -1.60 – -1.21 | **<0.001** | -0.72 | 0.11 | -0.94 – -0.50 | **<0.001** | 0.61 | 0.07 | 0.49 – 0.76 | **<0.001** | 0.21 | 0.02 | 0.17 – 0.25 | **<0.001** | 0.02 | 0.00 | 0.02 – 0.03 | **<0.001** | 0.01 | 0.00 | 0.00 – 0.01 | **<0.001** |
| Age | -0.01 | 0.00 | -0.02 – -0.01 | **<0.001** | -0.00 | 0.00 | -0.01 – 0.00 | 0.239 | 0.00 | 0.00 | -0.00 – 0.01 | 0.113 | 0.01 | 0.00 | 0.01 – 0.02 | **<0.001** | -0.01 | 0.00 | -0.01 – -0.00 | **0.002** | 0.01 | 0.00 | 0.00 – 0.01 | **<0.001** | 0.02 | 0.00 | 0.01 – 0.02 | **<0.001** | 0.01 | 0.00 | 0.01 – 0.02 | **<0.001** | 0.01 | 0.00 | 0.00 – 0.01 | **0.001** | 0.01 | 0.00 | 0.01 – 0.02 | **<0.001** | 0.03 | 0.00 | 0.03 – 0.04 | **<0.001** | 0.02 | 0.00 | 0.01 – 0.02 | **<0.001** | 0.98 | 0.00 | 0.98 – 0.99 | **<0.001** | 1.04 | 0.00 | 1.04 – 1.05 | **<0.001** | 1.04 | 0.00 | 1.03 – 1.04 | **<0.001** | 1.04 | 0.01 | 1.03 – 1.05 | **<0.001** |
| Sex | -0.01 | 0.18 | -0.36 – 0.34 | 0.970 | -1.65 | 0.15 | -1.95 – -1.36 | **<0.001** | -0.46 | 0.18 | -0.81 – -0.10 | **0.012** | 0.20 | 0.18 | -0.16 – 0.55 | 0.276 | -0.69 | 0.18 | -1.03 – -0.34 | **<0.001** | -0.80 | 0.17 | -1.13 – -0.46 | **<0.001** | 0.14 | 0.18 | -0.22 – 0.50 | 0.443 | -0.16 | 0.19 | -0.53 – 0.22 | 0.408 | -0.70 | 0.18 | -1.05 – -0.35 | **<0.001** | 0.85 | 0.18 | 0.50 – 1.20 | **<0.001** | 1.12 | 0.17 | 0.79 – 1.45 | **<0.001** | 0.37 | 0.18 | 0.01 – 0.73 | **0.047** | 0.19 | 0.05 | 0.12 – 0.30 | **<0.001** | 2.17 | 0.38 | 1.54 – 3.05 | **<0.001** | 0.74 | 0.25 | 0.38 – 1.43 | 0.379 | 1.95 | 1.00 | 0.70 – 5.29 | 0.195 |
| Inactivity (min) | 0.00 | 0.00 | -0.01 – 0.01 | 0.637 | 0.01 | 0.00 | 0.00 – 0.02 | **0.008** | 0.00 | 0.00 | -0.01 – 0.01 | 0.474 | 0.00 | 0.00 | -0.01 – 0.01 | 0.981 | 0.01 | 0.00 | -0.00 – 0.01 | 0.150 | 0.00 | 0.00 | -0.01 – 0.01 | 0.576 | -0.01 | 0.00 | -0.02 – -0.00 | **0.011** | -0.01 | 0.00 | -0.01 – 0.00 | 0.244 | 0.01 | 0.00 | -0.00 – 0.01 | 0.197 | -0.01 | 0.00 | -0.02 – -0.00 | **0.005** | -0.02 | 0.00 | -0.03 – -0.01 | **<0.001** | -0.01 | 0.00 | -0.02 – 0.00 | 0.080 | 1.01 | 0.01 | 1.00 – 1.02 | **0.045** | 0.99 | 0.00 | 0.98 – 1.00 | **0.002** | 1.00 | 0.01 | 0.98 – 1.01 | 0.725 | 0.98 | 0.01 | 0.96 – 1.00 | **0.038** |
| Age:Sex | 0.18 | 0.03 | 0.12 – 0.24 | **<0.001** | 0.17 | 0.03 | 0.11 – 0.22 | **<0.001** | 0.18 | 0.03 | 0.12 – 0.25 | **<0.001** | 0.07 | 0.03 | 0.01 – 0.14 | **0.026** | 0.17 | 0.03 | 0.11 – 0.23 | **<0.001** | 0.17 | 0.03 | 0.11 – 0.23 | **<0.001** | 0.00 | 0.03 | -0.06 – 0.07 | 0.975 | 0.00 | 0.03 | -0.06 – 0.07 | 0.921 | -0.05 | 0.03 | -0.11 – 0.02 | 0.138 | 0.13 | 0.03 | 0.06 – 0.19 | **<0.001** | 0.03 | 0.03 | -0.03 – 0.09 | 0.263 | 0.09 | 0.03 | 0.02 – 0.15 | **0.009** | 1.71 | 0.07 | 1.58 – 1.85 | **<0.001** | 1.17 | 0.04 | 1.10 – 1.24 | **<0.001** | 1.08 | 0.06 | 0.98 – 1.20 | 0.130 | 1.21 | 0.10 | 1.03 – 1.42 | **0.023** |
| Observations | 1056 | | | | 1033 | | | | 1017 | | | | 1012 | | | | 1056 | | | | 1017 | | | | 1020 | | | | 955 | | | | 1020 | | | | 1020 | | | | 1016 | | | | 1016 | | | | 1056 | | | | 1016 | | | | 1012 | | | | 1012 | | | |

### Physical activity predictor: Inactivity

|  | Weight | | | | Body fat | | | | Waist circumference | | | | Waist:hip ratio | | | | BMI | | | | BRI | | | | Total cholesterol | | | | LDL | | | | HDL | | | | Triglycerides | | | | Systolic BP | | | | Diastolic BP | | | | Obese (y/n) | | | | Hypertension (y/n) | | | | Pre-diabetes (y/n) | | | | Diabetes (y/n) | | | |
| --- | --- | --- | --- | --- | --- | --- | --- | --- | --- | --- | --- | --- | --- | --- | --- | --- | --- | --- | --- | --- | --- | --- | --- | --- | --- | --- | --- | --- | --- | --- | --- | --- | --- | --- | --- | --- | --- | --- | --- | --- | --- | --- | --- | --- | --- | --- | --- | --- | --- | --- | --- | --- | --- | --- | --- | --- | --- | --- | --- | --- | --- | --- | --- | --- |
| Predictors | Estimates | std. Error | CI | p | Estimates | std. Error | CI | p | Estimates | std. Error | CI | p | Estimates | std. Error | CI | p | Estimates | std. Error | CI | p | Estimates | std. Error | CI | p | Estimates | std. Error | CI | p | Estimates | std. Error | CI | p | Estimates | std. Error | CI | p | Estimates | std. Error | CI | p | Estimates | std. Error | CI | p | Estimates | std. Error | CI | p | Odds Ratios | std. Error | CI | p | Odds Ratios | std. Error | CI | p | Odds Ratios | std. Error | CI | p | Odds Ratios | std. Error | CI | p |
  
  

### Physical activity predictor: Intensity gradient

| Intercept | 0.42 | 0.11 | 0.21 – 0.63 | **<0.001** | 0.55 | 0.09 | 0.38 – 0.73 | **<0.001** | -0.08 | 0.11 | -0.29 – 0.14 | 0.482 | -0.66 | 0.11 | -0.88 – -0.45 | **<0.001** | 0.46 | 0.11 | 0.25 – 0.67 | **<0.001** | -0.14 | 0.10 | -0.34 – 0.07 | 0.189 | -0.50 | 0.11 | -0.72 – -0.28 | **<0.001** | -0.32 | 0.12 | -0.54 – -0.09 | **0.006** | -0.19 | 0.11 | -0.40 – 0.02 | 0.074 | -0.57 | 0.11 | -0.78 – -0.36 | **<0.001** | -1.41 | 0.10 | -1.60 – -1.21 | **<0.001** | -0.72 | 0.11 | -0.94 – -0.50 | **<0.001** | 0.61 | 0.07 | 0.49 – 0.76 | **<0.001** | 0.21 | 0.02 | 0.17 – 0.25 | **<0.001** | 0.02 | 0.00 | 0.02 – 0.03 | **<0.001** | 0.01 | 0.00 | 0.00 – 0.01 | **<0.001** |
| Age | -0.01 | 0.00 | -0.02 – -0.01 | **<0.001** | -0.00 | 0.00 | -0.01 – 0.00 | 0.239 | 0.00 | 0.00 | -0.00 – 0.01 | 0.113 | 0.01 | 0.00 | 0.01 – 0.02 | **<0.001** | -0.01 | 0.00 | -0.01 – -0.00 | **0.002** | 0.01 | 0.00 | 0.00 – 0.01 | **<0.001** | 0.02 | 0.00 | 0.01 – 0.02 | **<0.001** | 0.01 | 0.00 | 0.01 – 0.02 | **<0.001** | 0.01 | 0.00 | 0.00 – 0.01 | **0.001** | 0.01 | 0.00 | 0.01 – 0.02 | **<0.001** | 0.03 | 0.00 | 0.03 – 0.04 | **<0.001** | 0.02 | 0.00 | 0.01 – 0.02 | **<0.001** | 0.98 | 0.00 | 0.98 – 0.99 | **<0.001** | 1.04 | 0.00 | 1.04 – 1.05 | **<0.001** | 1.04 | 0.00 | 1.03 – 1.04 | **<0.001** | 1.04 | 0.01 | 1.03 – 1.05 | **<0.001** |
| Sex | -0.01 | 0.18 | -0.36 – 0.34 | 0.970 | -1.65 | 0.15 | -1.95 – -1.36 | **<0.001** | -0.46 | 0.18 | -0.81 – -0.10 | **0.012** | 0.20 | 0.18 | -0.16 – 0.55 | 0.276 | -0.69 | 0.18 | -1.03 – -0.34 | **<0.001** | -0.80 | 0.17 | -1.13 – -0.46 | **<0.001** | 0.14 | 0.18 | -0.22 – 0.50 | 0.443 | -0.16 | 0.19 | -0.53 – 0.22 | 0.408 | -0.70 | 0.18 | -1.05 – -0.35 | **<0.001** | 0.85 | 0.18 | 0.50 – 1.20 | **<0.001** | 1.12 | 0.17 | 0.79 – 1.45 | **<0.001** | 0.37 | 0.18 | 0.01 – 0.73 | **0.047** | 0.19 | 0.05 | 0.12 – 0.30 | **<0.001** | 2.17 | 0.38 | 1.54 – 3.05 | **<0.001** | 0.74 | 0.25 | 0.38 – 1.43 | 0.379 | 1.95 | 1.00 | 0.70 – 5.29 | 0.195 |
| Intensity gradient | 0.00 | 0.00 | -0.01 – 0.01 | 0.637 | 0.01 | 0.00 | 0.00 – 0.02 | **0.008** | 0.00 | 0.00 | -0.01 – 0.01 | 0.474 | 0.00 | 0.00 | -0.01 – 0.01 | 0.981 | 0.01 | 0.00 | -0.00 – 0.01 | 0.150 | 0.00 | 0.00 | -0.01 – 0.01 | 0.576 | -0.01 | 0.00 | -0.02 – -0.00 | **0.011** | -0.01 | 0.00 | -0.01 – 0.00 | 0.244 | 0.01 | 0.00 | -0.00 – 0.01 | 0.197 | -0.01 | 0.00 | -0.02 – -0.00 | **0.005** | -0.02 | 0.00 | -0.03 – -0.01 | **<0.001** | -0.01 | 0.00 | -0.02 – 0.00 | 0.080 | 1.01 | 0.01 | 1.00 – 1.02 | **0.045** | 0.99 | 0.00 | 0.98 – 1.00 | **0.002** | 1.00 | 0.01 | 0.98 – 1.01 | 0.725 | 0.98 | 0.01 | 0.96 – 1.00 | **0.038** |
| Age:Sex | 0.18 | 0.03 | 0.12 – 0.24 | **<0.001** | 0.17 | 0.03 | 0.11 – 0.22 | **<0.001** | 0.18 | 0.03 | 0.12 – 0.25 | **<0.001** | 0.07 | 0.03 | 0.01 – 0.14 | **0.026** | 0.17 | 0.03 | 0.11 – 0.23 | **<0.001** | 0.17 | 0.03 | 0.11 – 0.23 | **<0.001** | 0.00 | 0.03 | -0.06 – 0.07 | 0.975 | 0.00 | 0.03 | -0.06 – 0.07 | 0.921 | -0.05 | 0.03 | -0.11 – 0.02 | 0.138 | 0.13 | 0.03 | 0.06 – 0.19 | **<0.001** | 0.03 | 0.03 | -0.03 – 0.09 | 0.263 | 0.09 | 0.03 | 0.02 – 0.15 | **0.009** | 1.71 | 0.07 | 1.58 – 1.85 | **<0.001** | 1.17 | 0.04 | 1.10 – 1.24 | **<0.001** | 1.08 | 0.06 | 0.98 – 1.20 | 0.130 | 1.21 | 0.10 | 1.03 – 1.42 | **0.023** |
| Observations | 1056 | | | | 1033 | | | | 1017 | | | | 1012 | | | | 1056 | | | | 1017 | | | | 1020 | | | | 955 | | | | 1020 | | | | 1020 | | | | 1016 | | | | 1016 | | | | 1056 | | | | 1016 | | | | 1012 | | | | 1012 | | | |

  
  

### Physical activity predictor: Daily steps

|  | Weight | | | | Body fat | | | | Waist circumference | | | | Waist:hip ratio | | | | BMI | | | | BRI | | | | Total cholesterol | | | | LDL | | | | HDL | | | | Triglycerides | | | | Systolic BP | | | | Diastolic BP | | | | Obese (y/n) | | | | Hypertension (y/n) | | | | Pre-diabetes (y/n) | | | | Diabetes (y/n) | | | |
| --- | --- | --- | --- | --- | --- | --- | --- | --- | --- | --- | --- | --- | --- | --- | --- | --- | --- | --- | --- | --- | --- | --- | --- | --- | --- | --- | --- | --- | --- | --- | --- | --- | --- | --- | --- | --- | --- | --- | --- | --- | --- | --- | --- | --- | --- | --- | --- | --- | --- | --- | --- | --- | --- | --- | --- | --- | --- | --- | --- | --- | --- | --- | --- | --- |
| Predictors | Estimates | std. Error | CI | p | Estimates | std. Error | CI | p | Estimates | std. Error | CI | p | Estimates | std. Error | CI | p | Estimates | std. Error | CI | p | Estimates | std. Error | CI | p | Estimates | std. Error | CI | p | Estimates | std. Error | CI | p | Estimates | std. Error | CI | p | Estimates | std. Error | CI | p | Estimates | std. Error | CI | p | Estimates | std. Error | CI | p | Odds Ratios | std. Error | CI | p | Odds Ratios | std. Error | CI | p | Odds Ratios | std. Error | CI | p | Odds Ratios | std. Error | CI | p |
| Intercept | 0.42 | 0.11 | 0.21 – 0.63 | **<0.001** | 0.55 | 0.09 | 0.38 – 0.73 | **<0.001** | -0.08 | 0.11 | -0.29 – 0.14 | 0.482 | -0.66 | 0.11 | -0.88 – -0.45 | **<0.001** | 0.46 | 0.11 | 0.25 – 0.67 | **<0.001** | -0.14 | 0.10 | -0.34 – 0.07 | 0.189 | -0.50 | 0.11 | -0.72 – -0.28 | **<0.001** | -0.32 | 0.12 | -0.54 – -0.09 | **0.006** | -0.19 | 0.11 | -0.40 – 0.02 | 0.074 | -0.57 | 0.11 | -0.78 – -0.36 | **<0.001** | -1.41 | 0.10 | -1.60 – -1.21 | **<0.001** | -0.72 | 0.11 | -0.94 – -0.50 | **<0.001** | 0.61 | 0.07 | 0.49 – 0.76 | **<0.001** | 0.21 | 0.02 | 0.17 – 0.25 | **<0.001** | 0.02 | 0.00 | 0.02 – 0.03 | **<0.001** | 0.01 | 0.00 | 0.00 – 0.01 | **<0.001** |
| Age | -0.01 | 0.00 | -0.02 – -0.01 | **<0.001** | -0.00 | 0.00 | -0.01 – 0.00 | 0.239 | 0.00 | 0.00 | -0.00 – 0.01 | 0.113 | 0.01 | 0.00 | 0.01 – 0.02 | **<0.001** | -0.01 | 0.00 | -0.01 – -0.00 | **0.002** | 0.01 | 0.00 | 0.00 – 0.01 | **<0.001** | 0.02 | 0.00 | 0.01 – 0.02 | **<0.001** | 0.01 | 0.00 | 0.01 – 0.02 | **<0.001** | 0.01 | 0.00 | 0.00 – 0.01 | **0.001** | 0.01 | 0.00 | 0.01 – 0.02 | **<0.001** | 0.03 | 0.00 | 0.03 – 0.04 | **<0.001** | 0.02 | 0.00 | 0.01 – 0.02 | **<0.001** | 0.98 | 0.00 | 0.98 – 0.99 | **<0.001** | 1.04 | 0.00 | 1.04 – 1.05 | **<0.001** | 1.04 | 0.00 | 1.03 – 1.04 | **<0.001** | 1.04 | 0.01 | 1.03 – 1.05 | **<0.001** |
| Sex | -0.01 | 0.18 | -0.36 – 0.34 | 0.970 | -1.65 | 0.15 | -1.95 – -1.36 | **<0.001** | -0.46 | 0.18 | -0.81 – -0.10 | **0.012** | 0.20 | 0.18 | -0.16 – 0.55 | 0.276 | -0.69 | 0.18 | -1.03 – -0.34 | **<0.001** | -0.80 | 0.17 | -1.13 – -0.46 | **<0.001** | 0.14 | 0.18 | -0.22 – 0.50 | 0.443 | -0.16 | 0.19 | -0.53 – 0.22 | 0.408 | -0.70 | 0.18 | -1.05 – -0.35 | **<0.001** | 0.85 | 0.18 | 0.50 – 1.20 | **<0.001** | 1.12 | 0.17 | 0.79 – 1.45 | **<0.001** | 0.37 | 0.18 | 0.01 – 0.73 | **0.047** | 0.19 | 0.05 | 0.12 – 0.30 | **<0.001** | 2.17 | 0.38 | 1.54 – 3.05 | **<0.001** | 0.74 | 0.25 | 0.38 – 1.43 | 0.379 | 1.95 | 1.00 | 0.70 – 5.29 | 0.195 |
| Daily steps | 0.00 | 0.00 | -0.01 – 0.01 | 0.637 | 0.01 | 0.00 | 0.00 – 0.02 | **0.008** | 0.00 | 0.00 | -0.01 – 0.01 | 0.474 | 0.00 | 0.00 | -0.01 – 0.01 | 0.981 | 0.01 | 0.00 | -0.00 – 0.01 | 0.150 | 0.00 | 0.00 | -0.01 – 0.01 | 0.576 | -0.01 | 0.00 | -0.02 – -0.00 | **0.011** | -0.01 | 0.00 | -0.01 – 0.00 | 0.244 | 0.01 | 0.00 | -0.00 – 0.01 | 0.197 | -0.01 | 0.00 | -0.02 – -0.00 | **0.005** | -0.02 | 0.00 | -0.03 – -0.01 | **<0.001** | -0.01 | 0.00 | -0.02 – 0.00 | 0.080 | 1.01 | 0.01 | 1.00 – 1.02 | **0.045** | 0.99 | 0.00 | 0.98 – 1.00 | **0.002** | 1.00 | 0.01 | 0.98 – 1.01 | 0.725 | 0.98 | 0.01 | 0.96 – 1.00 | **0.038** |
| Age:Sex | 0.18 | 0.03 | 0.12 – 0.24 | **<0.001** | 0.17 | 0.03 | 0.11 – 0.22 | **<0.001** | 0.18 | 0.03 | 0.12 – 0.25 | **<0.001** | 0.07 | 0.03 | 0.01 – 0.14 | **0.026** | 0.17 | 0.03 | 0.11 – 0.23 | **<0.001** | 0.17 | 0.03 | 0.11 – 0.23 | **<0.001** | 0.00 | 0.03 | -0.06 – 0.07 | 0.975 | 0.00 | 0.03 | -0.06 – 0.07 | 0.921 | -0.05 | 0.03 | -0.11 – 0.02 | 0.138 | 0.13 | 0.03 | 0.06 – 0.19 | **<0.001** | 0.03 | 0.03 | -0.03 – 0.09 | 0.263 | 0.09 | 0.03 | 0.02 – 0.15 | **0.009** | 1.71 | 0.07 | 1.58 – 1.85 | **<0.001** | 1.17 | 0.04 | 1.10 – 1.24 | **<0.001** | 1.08 | 0.06 | 0.98 – 1.20 | 0.130 | 1.21 | 0.10 | 1.03 – 1.42 | **0.023** |
| Observations | 1056 | | | | 1033 | | | | 1017 | | | | 1012 | | | | 1056 | | | | 1017 | | | | 1020 | | | | 955 | | | | 1020 | | | | 1020 | | | | 1016 | | | | 1016 | | | | 1056 | | | | 1016 | | | | 1012 | | | | 1012 | | | |
