## Supplementary Table 5 for "Physical activity and cardiometabolic health across an extreme lifestyle gradient"

|  | Weight | | | | Body fat | | | | Waist circumference | | | | Waist:hip ratio | | | | BMI | | | | BRI | | | | Total cholesterol | | | | LDL | | | | HDL | | | | Triglycerides | | | | Systolic BP | | | | Diastolic BP | | | | Obese (y/n) | | | | Hypertension (y/n) | | | | Pre-diabetes (y/n) | | | | Diabetes (y/n) | | | |
| --- | --- | --- | --- | --- | --- | --- | --- | --- | --- | --- | --- | --- | --- | --- | --- | --- | --- | --- | --- | --- | --- | --- | --- | --- | --- | --- | --- | --- | --- | --- | --- | --- | --- | --- | --- | --- | --- | --- | --- | --- | --- | --- | --- | --- | --- | --- | --- | --- | --- | --- | --- | --- | --- | --- | --- | --- | --- | --- | --- | --- | --- | --- | --- | --- |
| Predictors | Estimates | std. Error | CI | p | Estimates | std. Error | CI | p | Estimates | std. Error | CI | p | Estimates | std. Error | CI | p | Estimates | std. Error | CI | p | Estimates | std. Error | CI | p | Estimates | std. Error | CI | p | Estimates | std. Error | CI | p | Estimates | std. Error | CI | p | Estimates | std. Error | CI | p | Estimates | std. Error | CI | p | Estimates | std. Error | CI | p | Odds Ratios | std. Error | CI | p | Odds Ratios | std. Error | CI | p | Odds Ratios | std. Error | CI | p | Odds Ratios | std. Error | CI | p |
| Intercept | 0.60 | 0.10 | 0.40 – 0.79 | **<0.001** | 0.67 | 0.09 | 0.50 – 0.84 | **<0.001** | 0.05 | 0.10 | -0.15 – 0.26 | 0.628 | -0.66 | 0.11 | -0.87 – -0.44 | **<0.001** | 0.62 | 0.10 | 0.42 – 0.82 | **<0.001** | -0.03 | 0.10 | -0.23 – 0.17 | 0.767 | -0.46 | 0.11 | -0.68 – -0.24 | **<0.001** | -0.30 | 0.12 | -0.52 – -0.07 | **0.011** | -0.16 | 0.11 | -0.38 – 0.05 | 0.130 | -0.50 | 0.11 | -0.71 – -0.29 | **<0.001** | -1.43 | 0.10 | -1.63 – -1.23 | **<0.001** | -0.74 | 0.11 | -0.96 – -0.52 | **<0.001** | 0.79 | 0.10 | 0.62 – 1.02 | 0.066 | 0.20 | 0.02 | 0.16 – 0.24 | **<0.001** | 0.02 | 0.00 | 0.02 – 0.03 | **<0.001** | 0.01 | 0.00 | 0.00 – 0.01 | **<0.001** |
| Urbanicity score | 0.41 | 0.03 | 0.35 – 0.47 | **<0.001** | 0.26 | 0.03 | 0.21 – 0.30 | **<0.001** | 0.32 | 0.03 | 0.26 – 0.38 | **<0.001** | 0.04 | 0.03 | -0.02 – 0.10 | 0.196 | 0.37 | 0.03 | 0.31 – 0.43 | **<0.001** | 0.27 | 0.03 | 0.21 – 0.33 | **<0.001** | 0.10 | 0.03 | 0.04 – 0.17 | **0.002** | 0.05 | 0.03 | -0.02 – 0.12 | 0.144 | 0.07 | 0.03 | 0.01 – 0.13 | **0.030** | 0.20 | 0.03 | 0.14 – 0.26 | **<0.001** | -0.07 | 0.03 | -0.13 – -0.01 | **0.023** | -0.03 | 0.03 | -0.09 – 0.04 | 0.395 | 2.43 | 0.10 | 2.24 – 2.64 | **<0.001** | 0.91 | 0.03 | 0.85 – 0.96 | **0.001** | 1.46 | 0.08 | 1.31 – 1.62 | **<0.001** | 1.49 | 0.13 | 1.25 – 1.77 | **<0.001** |
| Age | -0.02 | 0.00 | -0.02 – -0.01 | **<0.001** | -0.01 | 0.00 | -0.01 – -0.00 | **0.006** | 0.00 | 0.00 | -0.00 – 0.01 | 0.849 | 0.01 | 0.00 | 0.01 – 0.02 | **<0.001** | -0.01 | 0.00 | -0.02 – -0.01 | **<0.001** | 0.01 | 0.00 | 0.00 – 0.01 | **0.014** | 0.01 | 0.00 | 0.01 – 0.02 | **<0.001** | 0.01 | 0.00 | 0.01 – 0.02 | **<0.001** | 0.01 | 0.00 | 0.00 – 0.01 | **0.002** | 0.01 | 0.00 | 0.00 – 0.01 | **0.002** | 0.03 | 0.00 | 0.03 – 0.04 | **<0.001** | 0.02 | 0.00 | 0.01 – 0.02 | **<0.001** | 0.97 | 0.00 | 0.96 – 0.98 | **<0.001** | 1.04 | 0.00 | 1.04 – 1.05 | **<0.001** | 1.03 | 0.00 | 1.02 – 1.04 | **<0.001** | 1.04 | 0.01 | 1.03 – 1.05 | **<0.001** |
| Sex | 0.13 | 0.17 | -0.20 – 0.45 | 0.435 | -1.57 | 0.14 | -1.85 – -1.29 | **<0.001** | -0.34 | 0.17 | -0.68 – 0.00 | 0.051 | 0.22 | 0.18 | -0.13 – 0.57 | 0.218 | -0.57 | 0.17 | -0.89 – -0.24 | **0.001** | -0.70 | 0.17 | -1.03 – -0.37 | **<0.001** | 0.18 | 0.18 | -0.18 – 0.54 | 0.334 | -0.14 | 0.19 | -0.52 – 0.23 | 0.453 | -0.68 | 0.18 | -1.04 – -0.33 | **<0.001** | 0.94 | 0.18 | 0.59 – 1.28 | **<0.001** | 1.10 | 0.17 | 0.77 – 1.42 | **<0.001** | 0.37 | 0.18 | 0.01 – 0.73 | **0.046** | 0.19 | 0.05 | 0.11 – 0.33 | **<0.001** | 2.14 | 0.37 | 1.51 – 3.01 | **<0.001** | 0.85 | 0.29 | 0.43 – 1.65 | 0.628 | 2.32 | 1.22 | 0.81 – 6.45 | 0.110 |
| Age:Sex | 0.00 | 0.00 | -0.01 – 0.01 | 0.637 | 0.01 | 0.00 | 0.00 – 0.02 | **0.006** | 0.00 | 0.00 | -0.01 – 0.01 | 0.514 | -0.00 | 0.00 | -0.01 – 0.01 | 0.976 | 0.01 | 0.00 | -0.00 – 0.01 | 0.138 | 0.00 | 0.00 | -0.01 – 0.01 | 0.625 | -0.01 | 0.00 | -0.02 – -0.00 | **0.010** | -0.01 | 0.00 | -0.01 – 0.00 | 0.239 | 0.01 | 0.00 | -0.00 – 0.01 | 0.175 | -0.01 | 0.00 | -0.02 – -0.00 | **0.003** | -0.02 | 0.00 | -0.03 – -0.01 | **<0.001** | -0.01 | 0.00 | -0.02 – 0.00 | 0.061 | 1.01 | 0.01 | 1.00 – 1.03 | **0.020** | 0.99 | 0.00 | 0.98 – 1.00 | **0.002** | 1.00 | 0.01 | 0.98 – 1.01 | 0.681 | 0.98 | 0.01 | 0.96 – 1.00 | **0.032** |
| Inactivity (min) | 0.12 | 0.03 | 0.06 – 0.18 | **<0.001** | 0.12 | 0.03 | 0.07 – 0.18 | **<0.001** | 0.13 | 0.03 | 0.07 – 0.19 | **<0.001** | 0.07 | 0.03 | 0.00 – 0.13 | **0.049** | 0.11 | 0.03 | 0.05 – 0.17 | **<0.001** | 0.13 | 0.03 | 0.06 – 0.19 | **<0.001** | -0.01 | 0.03 | -0.08 – 0.05 | 0.679 | -0.00 | 0.04 | -0.07 – 0.07 | 0.929 | -0.06 | 0.03 | -0.13 – 0.01 | 0.070 | 0.10 | 0.03 | 0.04 – 0.16 | **0.002** | 0.04 | 0.03 | -0.02 – 0.10 | 0.165 | 0.09 | 0.03 | 0.02 – 0.15 | **0.009** | 1.56 | 0.07 | 1.44 – 1.70 | **<0.001** | 1.18 | 0.04 | 1.11 – 1.25 | **<0.001** | 1.04 | 0.06 | 0.93 – 1.15 | 0.510 | 1.16 | 0.10 | 0.98 – 1.36 | 0.088 |
| Observations | 1044 | | | | 1027 | | | | 1011 | | | | 1006 | | | | 1044 | | | | 1011 | | | | 1014 | | | | 949 | | | | 1014 | | | | 1014 | | | | 1010 | | | | 1010 | | | | 1044 | | | | 1010 | | | | 1006 | | | | 1006 | | | |

### Physical activity predictor: Inactivity

|  | Weight | | | | Body fat | | | | Waist circumference | | | | Waist:hip ratio | | | | BMI | | | | BRI | | | | Total cholesterol | | | | LDL | | | | HDL | | | | Triglycerides | | | | Systolic BP | | | | Diastolic BP | | | | Obese (y/n) | | | | Hypertension (y/n) | | | | Pre-diabetes (y/n) | | | | Diabetes (y/n) |
| --- | --- | --- | --- | --- | --- | --- | --- | --- | --- | --- | --- | --- | --- | --- | --- | --- | --- | --- | --- | --- | --- | --- | --- | --- | --- | --- | --- | --- | --- | --- | --- | --- | --- | --- | --- | --- | --- | --- | --- | --- | --- | --- | --- | --- | --- | --- | --- | --- | --- | --- | --- | --- | --- | --- | --- | --- | --- | --- | --- | --- | --- |
  
  

### Physical activity predictor: Intensity gradient

| Predictors | Estimates | std. Error | CI | p | Estimates | std. Error | CI | p | Estimates | std. Error | CI | p | Estimates | std. Error | CI | p | Estimates | std. Error | CI | p | Estimates | std. Error | CI | p | Estimates | std. Error | CI | p | Estimates | std. Error | CI | p | Estimates | std. Error | CI | p | Estimates | std. Error | CI | p | Estimates | std. Error | CI | p | Estimates | std. Error | CI | p | Odds Ratios | std. Error | CI | p | Odds Ratios | std. Error | CI | p | Odds Ratios | std. Error | CI | p | Odds Ratios | std. Error | CI | p |
| Intercept | 0.62 | 0.10 | 0.42 – 0.82 | **<0.001** | 0.69 | 0.09 | 0.52 – 0.86 | **<0.001** | 0.07 | 0.11 | -0.13 – 0.28 | 0.487 | -0.67 | 0.11 | -0.89 – -0.46 | **<0.001** | 0.63 | 0.10 | 0.43 – 0.83 | **<0.001** | -0.02 | 0.10 | -0.21 – 0.18 | 0.876 | -0.45 | 0.11 | -0.67 – -0.23 | **<0.001** | -0.27 | 0.12 | -0.50 – -0.05 | **0.019** | -0.17 | 0.11 | -0.39 – 0.04 | 0.110 | -0.50 | 0.11 | -0.71 – -0.29 | **<0.001** | -1.43 | 0.10 | -1.63 – -1.23 | **<0.001** | -0.70 | 0.11 | -0.92 – -0.48 | **<0.001** | 0.88 | 0.11 | 0.69 – 1.12 | 0.292 | 0.20 | 0.02 | 0.16 – 0.25 | **<0.001** | 0.03 | 0.01 | 0.02 – 0.04 | **<0.001** | 0.01 | 0.00 | 0.00 – 0.01 | **<0.001** |
| Urbanicity score | 0.42 | 0.03 | 0.37 – 0.48 | **<0.001** | 0.26 | 0.03 | 0.21 – 0.31 | **<0.001** | 0.33 | 0.03 | 0.27 – 0.39 | **<0.001** | 0.05 | 0.03 | -0.01 – 0.11 | 0.104 | 0.39 | 0.03 | 0.33 – 0.44 | **<0.001** | 0.28 | 0.03 | 0.22 – 0.34 | **<0.001** | 0.10 | 0.03 | 0.03 – 0.16 | **0.003** | 0.04 | 0.03 | -0.03 – 0.10 | 0.301 | 0.07 | 0.03 | 0.01 – 0.13 | **0.033** | 0.20 | 0.03 | 0.14 – 0.27 | **<0.001** | -0.06 | 0.03 | -0.12 – -0.00 | **0.035** | -0.02 | 0.03 | -0.08 – 0.04 | 0.552 | 2.50 | 0.10 | 2.30 – 2.71 | **<0.001** | 0.93 | 0.03 | 0.88 – 0.98 | **0.013** | 1.43 | 0.08 | 1.29 – 1.59 | **<0.001** | 1.43 | 0.13 | 1.21 – 1.71 | **<0.001** |
| Age | -0.02 | 0.00 | -0.02 – -0.01 | **<0.001** | -0.01 | 0.00 | -0.01 – -0.00 | **0.001** | -0.00 | 0.00 | -0.01 – 0.00 | 0.751 | 0.01 | 0.00 | 0.01 – 0.02 | **<0.001** | -0.01 | 0.00 | -0.02 – -0.01 | **<0.001** | 0.01 | 0.00 | 0.00 – 0.01 | **0.049** | 0.01 | 0.00 | 0.01 – 0.02 | **<0.001** | 0.01 | 0.00 | 0.00 – 0.02 | **0.001** | 0.01 | 0.00 | 0.00 – 0.01 | **0.001** | 0.01 | 0.00 | 0.00 – 0.01 | **0.004** | 0.03 | 0.00 | 0.03 – 0.04 | **<0.001** | 0.02 | 0.00 | 0.01 – 0.02 | **<0.001** | 0.97 | 0.00 | 0.96 – 0.97 | **<0.001** | 1.04 | 0.00 | 1.04 – 1.05 | **<0.001** | 1.03 | 0.00 | 1.02 – 1.04 | **<0.001** | 1.03 | 0.01 | 1.02 – 1.05 | **<0.001** |
| Sex | 0.20 | 0.17 | -0.14 – 0.54 | 0.249 | -1.45 | 0.15 | -1.74 – -1.16 | **<0.001** | -0.23 | 0.18 | -0.58 – 0.13 | 0.211 | 0.21 | 0.19 | -0.16 – 0.57 | 0.267 | -0.51 | 0.17 | -0.85 – -0.17 | **0.003** | -0.59 | 0.17 | -0.93 – -0.24 | **0.001** | 0.18 | 0.19 | -0.19 – 0.56 | 0.336 | -0.08 | 0.20 | -0.47 – 0.31 | 0.689 | -0.75 | 0.19 | -1.12 – -0.39 | **<0.001** | 0.97 | 0.18 | 0.61 – 1.32 | **<0.001** | 1.14 | 0.17 | 0.80 – 1.48 | **<0.001** | 0.46 | 0.19 | 0.09 – 0.84 | **0.016** | 0.28 | 0.08 | 0.16 – 0.48 | **<0.001** | 2.62 | 0.48 | 1.84 – 3.75 | **<0.001** | 0.98 | 0.34 | 0.49 – 1.94 | 0.965 | 3.19 | 1.70 | 1.10 – 8.98 | **0.030** |
| Age:Sex | 0.00 | 0.00 | -0.01 – 0.01 | 0.614 | 0.01 | 0.00 | 0.00 – 0.02 | **0.010** | 0.00 | 0.00 | -0.01 – 0.01 | 0.596 | 0.00 | 0.00 | -0.01 – 0.01 | 0.912 | 0.01 | 0.00 | -0.00 – 0.01 | 0.126 | 0.00 | 0.00 | -0.01 – 0.01 | 0.732 | -0.01 | 0.00 | -0.02 – -0.00 | **0.008** | -0.01 | 0.00 | -0.01 – 0.00 | 0.176 | 0.01 | 0.00 | -0.00 – 0.01 | 0.156 | -0.01 | 0.00 | -0.02 – -0.00 | **0.004** | -0.02 | 0.00 | -0.03 – -0.01 | **<0.001** | -0.01 | 0.00 | -0.02 – 0.00 | 0.059 | 1.01 | 0.01 | 1.00 – 1.03 | **0.034** | 0.99 | 0.00 | 0.98 – 0.99 | **0.001** | 1.00 | 0.01 | 0.98 – 1.01 | 0.587 | 0.98 | 0.01 | 0.96 – 1.00 | **0.019** |
| Intensity gradient | -0.07 | 0.04 | -0.14 – -0.00 | **0.050** | -0.10 | 0.03 | -0.16 – -0.03 | **0.002** | -0.09 | 0.04 | -0.17 – -0.02 | **0.014** | 0.01 | 0.04 | -0.07 – 0.08 | 0.897 | -0.06 | 0.04 | -0.13 – 0.01 | 0.105 | -0.09 | 0.04 | -0.16 – -0.02 | **0.013** | -0.00 | 0.04 | -0.08 – 0.08 | 0.977 | -0.04 | 0.04 | -0.12 – 0.04 | 0.320 | 0.06 | 0.04 | -0.02 – 0.14 | 0.136 | -0.04 | 0.04 | -0.11 – 0.04 | 0.312 | -0.04 | 0.04 | -0.11 – 0.03 | 0.260 | -0.09 | 0.04 | -0.17 – -0.01 | **0.024** | 0.71 | 0.04 | 0.64 – 0.79 | **<0.001** | 0.87 | 0.03 | 0.80 – 0.93 | **<0.001** | 0.82 | 0.05 | 0.72 – 0.93 | **0.001** | 0.74 | 0.07 | 0.61 – 0.89 | **0.001** |
| Observations | 1059 | | | | 1041 | | | | 1026 | | | | 1021 | | | | 1059 | | | | 1026 | | | | 1029 | | | | 964 | | | | 1029 | | | | 1029 | | | | 1025 | | | | 1025 | | | | 1059 | | | | 1025 | | | | 1020 | | | | 1020 | | | |

  
  

### Physical activity predictor: Daily steps

|  | Weight | | | | Body fat | | | | Waist circumference | | | | Waist:hip ratio | | | | BMI | | | | BRI | | | | Total cholesterol | | | | LDL | | | | HDL | | | | Triglycerides | | | | Systolic BP | | | | Diastolic BP | | | | Obese (y/n) | | | | Hypertension (y/n) | | | | Pre-diabetes (y/n) | | | | Diabetes (y/n) | | | |
| --- | --- | --- | --- | --- | --- | --- | --- | --- | --- | --- | --- | --- | --- | --- | --- | --- | --- | --- | --- | --- | --- | --- | --- | --- | --- | --- | --- | --- | --- | --- | --- | --- | --- | --- | --- | --- | --- | --- | --- | --- | --- | --- | --- | --- | --- | --- | --- | --- | --- | --- | --- | --- | --- | --- | --- | --- | --- | --- | --- | --- | --- | --- | --- | --- |
| Predictors | Estimates | std. Error | CI | p | Estimates | std. Error | CI | p | Estimates | std. Error | CI | p | Estimates | std. Error | CI | p | Estimates | std. Error | CI | p | Estimates | std. Error | CI | p | Estimates | std. Error | CI | p | Estimates | std. Error | CI | p | Estimates | std. Error | CI | p | Estimates | std. Error | CI | p | Estimates | std. Error | CI | p | Estimates | std. Error | CI | p | Odds Ratios | std. Error | CI | p | Odds Ratios | std. Error | CI | p | Odds Ratios | std. Error | CI | p | Odds Ratios | std. Error | CI | p |
| Intercept | 0.62 | 0.10 | 0.42 – 0.81 | **<0.001** | 0.67 | 0.09 | 0.50 – 0.84 | **<0.001** | 0.05 | 0.10 | -0.15 – 0.26 | 0.611 | -0.66 | 0.11 | -0.87 – -0.45 | **<0.001** | 0.63 | 0.10 | 0.44 – 0.82 | **<0.001** | -0.04 | 0.10 | -0.23 – 0.16 | 0.704 | -0.44 | 0.11 | -0.65 – -0.22 | **<0.001** | -0.28 | 0.11 | -0.51 – -0.06 | **0.014** | -0.16 | 0.11 | -0.37 – 0.05 | 0.145 | -0.49 | 0.10 | -0.70 – -0.29 | **<0.001** | -1.44 | 0.10 | -1.63 – -1.24 | **<0.001** | -0.73 | 0.11 | -0.94 – -0.51 | **<0.001** | 0.80 | 0.10 | 0.63 – 1.02 | 0.068 | 0.19 | 0.02 | 0.16 – 0.24 | **<0.001** | 0.03 | 0.00 | 0.02 – 0.04 | **<0.001** | 0.01 | 0.00 | 0.00 – 0.01 | **<0.001** |
| Urbanicity score | 0.42 | 0.03 | 0.36 – 0.48 | **<0.001** | 0.26 | 0.03 | 0.21 – 0.31 | **<0.001** | 0.33 | 0.03 | 0.27 – 0.39 | **<0.001** | 0.05 | 0.03 | -0.02 – 0.11 | 0.152 | 0.38 | 0.03 | 0.32 – 0.44 | **<0.001** | 0.28 | 0.03 | 0.22 – 0.34 | **<0.001** | 0.09 | 0.03 | 0.03 – 0.15 | **0.005** | 0.03 | 0.03 | -0.03 – 0.10 | 0.316 | 0.07 | 0.03 | 0.00 – 0.13 | **0.036** | 0.20 | 0.03 | 0.14 – 0.26 | **<0.001** | -0.06 | 0.03 | -0.12 – -0.00 | **0.036** | -0.02 | 0.03 | -0.08 – 0.05 | 0.594 | 2.48 | 0.10 | 2.28 – 2.69 | **<0.001** | 0.92 | 0.03 | 0.87 – 0.98 | **0.008** | 1.45 | 0.08 | 1.31 – 1.62 | **<0.001** | 1.47 | 0.13 | 1.24 – 1.75 | **<0.001** |
| Age | -0.02 | 0.00 | -0.02 – -0.01 | **<0.001** | -0.01 | 0.00 | -0.01 – -0.00 | **0.003** | 0.00 | 0.00 | -0.00 – 0.01 | 0.939 | 0.01 | 0.00 | 0.01 – 0.02 | **<0.001** | -0.01 | 0.00 | -0.02 – -0.01 | **<0.001** | 0.01 | 0.00 | 0.00 – 0.01 | **0.014** | 0.01 | 0.00 | 0.01 – 0.02 | **<0.001** | 0.01 | 0.00 | 0.00 – 0.02 | **<0.001** | 0.01 | 0.00 | 0.00 – 0.01 | **0.002** | 0.01 | 0.00 | 0.00 – 0.01 | **0.002** | 0.03 | 0.00 | 0.03 – 0.04 | **<0.001** | 0.02 | 0.00 | 0.01 – 0.02 | **<0.001** | 0.97 | 0.00 | 0.96 – 0.98 | **<0.001** | 1.04 | 0.00 | 1.04 – 1.05 | **<0.001** | 1.03 | 0.00 | 1.02 – 1.04 | **<0.001** | 1.04 | 0.01 | 1.02 – 1.05 | **<0.001** |
| Sex | 0.17 | 0.17 | -0.16 – 0.49 | 0.310 | -1.51 | 0.14 | -1.79 – -1.23 | **<0.001** | -0.29 | 0.17 | -0.63 – 0.05 | 0.095 | 0.25 | 0.18 | -0.10 – 0.60 | 0.168 | -0.53 | 0.17 | -0.85 – -0.21 | **0.001** | -0.65 | 0.17 | -0.98 – -0.33 | **<0.001** | 0.21 | 0.18 | -0.15 – 0.57 | 0.249 | -0.08 | 0.19 | -0.46 – 0.29 | 0.659 | -0.70 | 0.18 | -1.05 – -0.35 | **<0.001** | 0.97 | 0.18 | 0.63 – 1.31 | **<0.001** | 1.11 | 0.17 | 0.78 – 1.43 | **<0.001** | 0.38 | 0.18 | 0.02 – 0.74 | **0.038** | 0.23 | 0.06 | 0.14 – 0.39 | **<0.001** | 2.40 | 0.42 | 1.70 – 3.38 | **<0.001** | 0.78 | 0.27 | 0.39 – 1.51 | 0.466 | 2.34 | 1.23 | 0.82 – 6.49 | 0.106 |
| Age:Sex | 0.00 | 0.00 | -0.01 – 0.01 | 0.633 | 0.01 | 0.00 | 0.00 – 0.02 | **0.008** | 0.00 | 0.00 | -0.01 – 0.01 | 0.544 | -0.00 | 0.00 | -0.01 – 0.01 | 0.945 | 0.01 | 0.00 | -0.00 – 0.01 | 0.133 | 0.00 | 0.00 | -0.01 – 0.01 | 0.657 | -0.01 | 0.00 | -0.02 – -0.00 | **0.005** | -0.01 | 0.00 | -0.01 – 0.00 | 0.153 | 0.01 | 0.00 | -0.00 – 0.01 | 0.183 | -0.01 | 0.00 | -0.02 – -0.00 | **0.003** | -0.02 | 0.00 | -0.03 – -0.01 | **<0.001** | -0.01 | 0.00 | -0.02 – 0.00 | 0.079 | 1.01 | 0.01 | 1.00 – 1.03 | **0.031** | 0.99 | 0.00 | 0.98 – 0.99 | **0.001** | 1.00 | 0.01 | 0.99 – 1.01 | 0.879 | 0.98 | 0.01 | 0.96 – 1.00 | **0.041** |
| Daily steps | -0.11 | 0.03 | -0.16 – -0.05 | **<0.001** | -0.11 | 0.03 | -0.16 – -0.06 | **<0.001** | -0.10 | 0.03 | -0.16 – -0.04 | **0.001** | -0.05 | 0.03 | -0.11 – 0.01 | 0.124 | -0.09 | 0.03 | -0.15 – -0.04 | **0.001** | -0.09 | 0.03 | -0.15 – -0.03 | **0.002** | -0.05 | 0.03 | -0.11 – 0.02 | 0.162 | -0.06 | 0.03 | -0.13 – 0.00 | 0.053 | 0.04 | 0.03 | -0.02 – 0.11 | 0.164 | -0.09 | 0.03 | -0.15 – -0.03 | **0.005** | -0.04 | 0.03 | -0.09 – 0.02 | 0.223 | -0.07 | 0.03 | -0.13 – -0.00 | **0.039** | 0.66 | 0.03 | 0.61 – 0.72 | **<0.001** | 0.84 | 0.03 | 0.79 – 0.89 | **<0.001** | 0.95 | 0.05 | 0.86 – 1.05 | 0.343 | 0.89 | 0.08 | 0.75 – 1.05 | 0.156 |
| Observations | 1059 | | | | 1041 | | | | 1026 | | | | 1021 | | | | 1059 | | | | 1026 | | | | 1029 | | | | 964 | | | | 1029 | | | | 1029 | | | | 1025 | | | | 1025 | | | | 1059 | | | | 1025 | | | | 1020 | | | | 1020 | | | |
